# ACK1 and BRK non-receptor tyrosine kinase deficiencies are associated with familial systemic lupus and involved in efferocytosis

**DOI:** 10.1101/2024.02.15.24302255

**Authors:** Stephanie Guillet, Tomi Lazarov, Natasha Jordan, Bertrand Boisson, Maria Tello, Barbara Craddock, Ting Zhou, Chihiro Nishi, Rohan Bareja, Hairu Yang, Frederic Rieux-Laucat, Rosa Irene Fregel Lorenzo, Sabrina D. Dyall, David Isenberg, David D’Cruz, Nico Lachmann, Olivier Elemento, Agnes Viale, Nicholas D. Socci, Laurent Abel, Shigekazu Nagata, Morgan Huse, W. Todd Miller, Jean-Laurent Casanova, Frederic Geissmann

**Author notes:** SG and TL equally contributed to this study. J-LC and W-TM equally contributed to this study. Correspondence to Frederic Geissmann.

## Abstract

Systemic Lupus Erythematosus (SLE) is an autoimmune disease, the pathophysiology and genetic basis of which are incompletely understood. Using a forward genetic screen in multiplex families with systemic lupus erythematosus (SLE) we identified an association between SLE and compound heterozygous deleterious variants in the non-receptor tyrosine kinases (NRTKs) ACK1 and BRK. Experimental blockade of ACK1 or BRK increased circulating autoantibodies *in vivo* in mice and exacerbated glomerular IgG deposits in an SLE mouse model. Mechanistically, non-receptor tyrosine kinases (NRTKs) regulate activation, migration, and proliferation of immune cells. We found that the patients’ ACK1 and BRK variants impair efferocytosis, the MERTK-mediated anti-inflammatory response to apoptotic cells, in human induced Pluripotent Stem Cell (hiPSC)-derived macrophages, which may contribute to SLE pathogenesis. Overall, our data suggest that ACK1 and BRK deficiencies are associated with human SLE and impair efferocytosis in macrophages.

**One sentence summary:** Loss of function variants of human ACK1 and BRK kinase underlie systemic lupus erythematosus in young patients from multiplex families and disrupt the anti-inflammatory response of macrophages to apoptotic cells.

## INTRODUCTION

Systemic lupus erythematosus (SLE) is a chronic autoimmune rheumatic disease, characterized by the presence of circulating autoantibodies against nuclear antigens. Clinical manifestations vary among affected individuals and can involve many organs and systems, including the skin, joints, kidneys, heart, lungs, nervous system, and hematopoietic system ^1,2^. The prevalence of SLE ranges from 0.4 to 2 /1,000, and varies with sex, age, and ancestry, being more common in women of childbearing age and individuals of African, Asian, and Hispanic ancestry ^1–9^. Sex, hormones, and environmental factors, including drugs and chemical exposures, viral infections, and sunlight, contribute to disease. At present specific therapies for SLE are few and far between ^10^, and clinical manifestations such as lupus nephritis, one of the most common and serious manifestations of SLE, remain a major risk factor for morbidity and mortality ^11–14^. The contribution of genetics to SLE is supported by epidemiological data showing familial aggregation ^15–17^ and higher concordance rates between monozygotic than dizygotic twins ^18^, the association of autosomal recessive deficiency in PKCdelta or DNAse1L3 with familial SLE, and similar phenotypes in the corresponding mouse models ^19–24^. In addition, genome wide association studies in large populations of patients have implicated a number of genes associated with immune system function ^1,25^, and an SLE-like disease is observed in a proportion of patients with immunodeficiency due to autosomal recessive or X-linked C1q, C1r/s, C2, C4, and NADPH-Oxidase deficiencies ^26–30^, with Rasopathies due to autosomal dominant gain-of-function mutations in the RAS pathway ^31^, and a proportion of patients presenting with interferonopathies with bi-allelic or mono-allelic mutations in genes coding for nucleic acid sensors such as TREX1, STING, SAMHD1, ADAR, and IFIH1 ^32–35^.

In this study we performed a forward genetic screen in multiplex SLE families with a well-defined phenotype, lupus nephritis, to identify new SLE causing genes and molecular pathways involved in SLE. We report the characterization of novel and rare deleterious variant alleles of two genes encoding the non-receptor tyrosine kinases (NRTK) Tyrosine Kinase Non-Receptor 2 / Activated CDC42 kinase 1 (TNK2/ACK1) and Protein Tyrosine Kinase 6 / Breast Tumor Kinase (PTK6/BRK) in two multiplex families. The variant alleles strongly decrease ACK1 and BRK kinase activity. ACK1 and BRK were shown to control B-cell and T-cell proliferation, survival, and activation ^36,37^. Although ACK1- and BRK-genetic deficiency ^37,38^ or their pharmacological inhibition does not result in spontaneous lupus development in C57BL/6 mice, we show that ACK1 or BRK inhibitors aggravate IgG glomerular deposition in the kidneys of BALB/cByJ mice treated with pristane to induce a lupus-like disease ^39^ and increases serum autoantibody levels. NRTKs mediate phosphorylation of downstream effectors, including RAC1, AKT, and STAT1/3, which are involved in immune cells homeostasis, and their deficiency can cause autoimmunity through different mechanisms ^40^. NRTKs such as ACK1 ^41^, Src ^42^, and PTK2/FAK ^43,44^ are also targets of MERTK, which mediates efferocytosis, the recognition of phosphatidylserine (PtdSer) on apoptotic cells for their anti-inflammatory engulfment ^45–48^. MERTK deficiency is a cause of SLE-like disease ^48^. We found that the patients’ ACK1 and BRK variants are kinase dead in response to MERTK activation and fail to phosphorylate AKT and STAT3, and to activate RAC1. Human iPSC-derived macrophages from patients and isogenic variants presented with defective AKT/STAT3 driven anti-inflammatory response and control of TNF and IL1β production in response to apoptotic cells and had a modest decrease in uptake of apoptotic cells, in comparison to familial and isogenic controls. In contrast, ACK1 and BRK kinase activity are dispensable for the phagocytosis of polystyrene beads, opsonized cells, and microbes. These results altogether suggest that ACK1 and BRK deficiencies underlie SLE in the two families, and that a defective efferocytic response to apoptotic cells may contribute to the auto-immune phenotype of the patients.

## RESULTS

### NRTK compound heterozygous missense variants in two multiplex families with SLE

We recruited 10 multiplex SLE families, each with 2 or 3 individuals diagnosed with biopsy confirmed lupus nephritis, classified according to the SLICC criteria ^2^ for whom we obtained genomic DNA (blood) in the Louise Coote Lupus Clinic at Guy’s and St Thomas’ Hospitals, London. We collected peripheral blood from a total of 22 patients and 17 relatives. Thirty five percent (%) of patients were male, and the same proportion were diagnosed before the age of 18 years. Genomic DNA was submitted to whole exome sequencing. Polymorphisms with a minor allele frequency (MAF) >0.01 in the publicly available database gnomAD (120,000 individuals), 1,000 genome Project (2,504 individuals) and our in-house database (>10,000 individuals) were excluded from analysis. We analyzed each kindred independently, under X-linked recessive, autosomal dominant, or autosomal recessive models of inheritance, and this analysis identified candidate genes in two kindreds.

In Family 1, we identified compound heterozygous missense variants in the non-receptor tyrosine kinase (NRTK) ACK1, which were confirmed by Sanger sequencing (**Figure 1A**, **Figure 1-figure supplement 1**). The 2 patients were males and developed a class IV lupus nephritis between age 10-15. The K161Q allele was inherited from one parent and A156T from the other (**Figure 1A**). Principal components analysis (PCA) based on the whole-exome sequencing to analyze population structure, parental inbreeding, and familial linkage ^49^ reveals South Asian ancestry, the closest 1,000 Genomes Project individuals being those from North India (India, Bangladesh, and Pakistan, see Methods). The ACK1 mutant alleles, A156T and K161Q (transcript ENST00000333602, **Figure 1-figure supplement 1**), have not been reported in South Asian (31,442 alleles) or other populations from public database (gnomAD, 1,000 genomes project) or our own in-house databases of >10,000 exomes of patients with infectious phenotypes. Finally, these variants were not found in DNA from 100 individuals from the small southeastern island from which they originated (see Methods). Altogether, ACK1 mutant alleles, A156T and K161Q are private to this family and their segregation is compatible with an autosomal recessive trait with complete penetrance.

**Figure 1.**
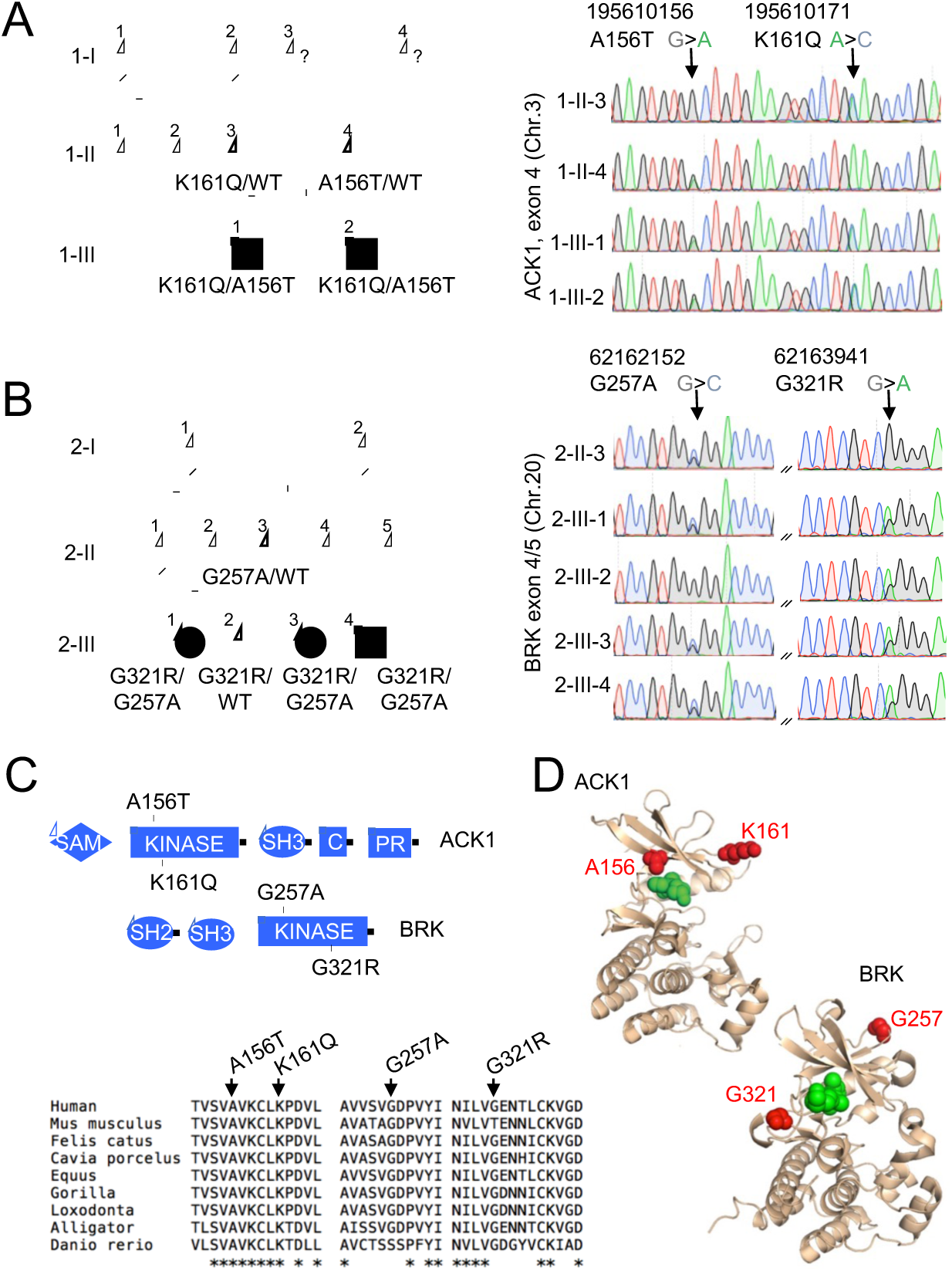
NRTK compound heterozygous missense variants in two multiplex families with SLE. (**A, B**) **Pedigrees and Sanger re-sequencing** of DNA from patients and healthy relatives of kindred 1 (A) carrying K161Q and A156T ACK1 mutations and kindred 2 (B) carrying G257A and G321R BRK mutations. Individuals with SLE are indicated by black boxes; deceased individuals are shown by diagonal bar; bold indicates the members analyzed by WES; squares indicate males, circles indicate females, and hexagons indicate generation I or II individuals with undisclosed sex for confidentiality. Black: Guanine, green: Adenine, red: Thymidine, blue: Cytosine. Arrows indicate nucleotide substitutions. Text indicates amino-acid substitutions. **(C) Domain architecture** (top panel) of ACK1 and BRK, with indicated mutations. SH2, Src homology 2; SH3, Src homology 3; Kinase, tyrosine kinase domain; C, Cdc42 binding domain; PR, Proline rich domain; SAM, Sterile α motif. **Alignment of kinase domains** (bottom panel) from ACK1 and BRK orthologs. Arrows indicate positions of mutations and stars indicate the amino acids conserved throughout species. **(D) Three dimensional (3D) structures of ACK1 and BRK.** *Top*: the crystal structure of ACK1 in a complex with AMP-PCP (PDB ID: 1U54). The mutated residues (A156 and K161) are shown in red, and the nucleotide analog is in green. *Bottom*: the crystal structure of BRK in a complex with the ATP-competitive inhibitor dasatinib (PDB ID: 5H2U). The mutated residues (G257 and G321) are shown in red, and dasatinib is in green.

In Family 2, we identified compound heterozygous missense variants in another NRTK, BRK in three siblings (**Figure 1B**, **Figure 1-figure supplement 1**). Two patients developed lupus nephritis, and the third patient developed a severe panniculitis, between the ages of 20 and 30 years. Two patients were female and one male. One parent, who was not genotyped, probably transmitted the G321R to their children, and the 2 other parents (who are also siblings) transmitted the G257A allele (only one parent was available for genotyping) (**Figure 1B**, **Figure 1-figure supplement 2**). An unaffected sibling (2.III.2) carries the G321R allele but not the G257A allele (**Figure 1B**, **Figure 1-figure supplement 2**). PCA analysis showed that individuals from family 2 have Sub-Saharan African ancestry, with the closest populations of 1,000 genomes project being African Caribbean in Barbados and African Ancestry in Southwest US and Luhya in Webuye in Kenya (see Methods). These 2 BRK mutant alleles, G257A and G321R are reported with a maximum MAF of ~ 8×10^−5^ and 5×10^−3^ respectively in Sub-Saharan African subpopulations (**Figure 1-figure supplement 1**), predicting a homozygosity frequency of ~ 10^−8^ and 10^−6^ respectively. These alleles are extremely rare outside Africa in the non-African gnomAD populations. These results are fully consistent under a recessive model with the overall prevalence of SLE (40 to 200 per 100,000 individuals). Genes that may cause SLE with high or low penetrance in accordance with their inheritance mode (**Figure 1-figure supplement 3**) were not candidates in these kindreds.

### The patients’ ACK1 and BRK variants are kinase null and hypomorphic

The ACK1 K161Q A156T and BRK G257A and G321R variants are all localized in the evolutionarily conserved kinase domains of the 2 proteins (**Figure 1C**), within the N-terminal (ATP-binding) lobes of the kinase catalytic domains ^50,51^ (**Figure 1D**), and near the positions of other mutations that decrease kinase activity ^52,53^ (**Figure 1C**). The variants are all predicted to be deleterious based on Combined Annotation Dependent Depletion (CADD) corrected with mutation significance cutoffs (MSC) ^54,55^ (**Figure 1-figure supplement 1**). To examine the functional effects of the mutations, we first expressed mutant forms of ACK1 and BRK in HEK293T cells. *In vitro* kinase assays indicated that ACK1 A156T and BRK G321R are kinase-dead mutants, and that ACK1 K161Q and BRK G257A are severe hypomorphs with a ~ *20%* residual kinase activity *in vitro* (**Figure 2A**). ACK1 A156T and K161Q and BRK G321R variants also lacked auto-phosphorylation activity (**Figure 2B**), phenocopying the effect of the specific ACK1 and BRK kinase inhibitors Aim100 ^56^ and Cpd4f ^57^ respectively (**Figure 2C**), while the BRK G257A allele has a small residual activity (**Figure 2C**). To examine the effects of the mutations in the patient’s cells we generated induced pluripotent stem cells (iPSC) from unrelated WT donors, patient 1-III-1 (ACK1 mutant), his heterozygous parent 1-II-3, patient 2-III-3 (BRK mutant) and her heterozygous sibling 2-III-2 (**Figure 2-figure supplement 1A-D**), and differentiated them into iPSC-derived macrophages ^58^ (**Figure 2D**, **Figure 2-figure supplement 1E**). hiPSC-derived macrophages from unrelated donors, familial controls, and patients presented with a normal morphology, survival, and phenotype and expressed comparable amount of ACK1 and BRK at the transcript and protein level (**Figure 2D, E**, **Figure 2-figure supplement 1G**), but an *in vitro* kinase assay indicated a loss of ACK1 kinase activity in ACK1^K161Q/A156T^ macrophages in comparison to controls (**Figure 2E**). The peptide substrate specificity of BRK overlaps with those of Src family kinases ^59^ present in the anti-BRK IPs. We therefore studied BRK kinase activity by IP-WB, which showed reduced BRK Tyr342 phosphorylation in BRK^G257A/G321R^ macrophages in comparison to controls (**Figure 2E**). These data altogether indicate that the patients’ ACK1 A156T and BRK G321R variants are kinase-dead alleles and K161Q and BRK G257A are severe hypomorphs.

**Figure 2.**
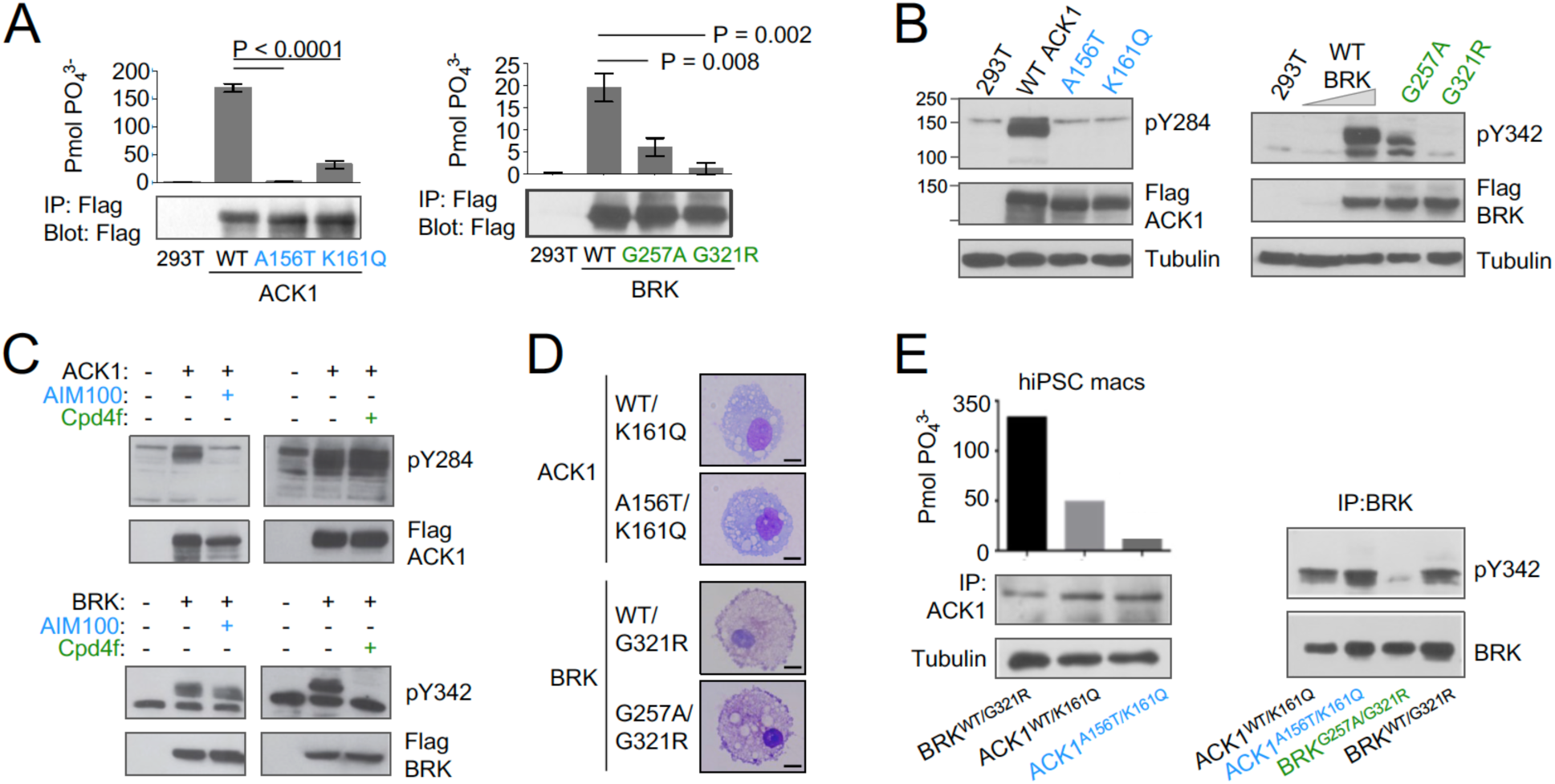
ACK1 and BRK mutations are null and hypomorph alleles. **(A) Immunoprecipitation (IP) kinase assay**. ACK1 *(Top)* was immunoprecipitated from 293T cells expressing Flag-tagged ACK1 wild type (WT), ACK1 A156T, or ACK1 K161Q with anti-Flag Ab. Immunoprecipitated proteins were used for duplicate in vitro kinase reactions with WASP synthetic peptide. Samples of the immunoprecipitates were also probed with anti-Flag Ab. BRK *(bottom)* was immunoprecipitated as above from 293T cells expressing Flag-tagged BRK WT and mutants with anti-Flag Ab. Kinase reactions were performed with peptide AEEEIYGEFEAKKKG, and represented as above, and samples of the immunoprecipitates probed with anti-Flag Ab. p-values were calculated using an *Anova* test (Tukey’s multiple comparisons test). **(B) Western blot analyses** of lysates from 293T cells expressing (left panel) Flag-tagged WT or mutant forms ACK1 probed with anti-ACK1 Tyr(P)^284^ (PY284), anti-Flag and anti-tubulin antibodies (Ab), and expressing (right panel) Flag-tagged WT or mutant BRK probed with anti-BRK Tyr(P)^342^ (PY342), anti-Flag and anti-tubulin antibodies. For BRK, 293T cells were starved overnight, and stimulated with 100 ng/ml EGF for 10 min. The lysate from WT BRK indicated as low was from cells transfected with one-tenth the amount of WT DNA. **(C)** Western blot analyses of lysates from 293T cells expressing ACK1-Flag or BRK-Flag treated with AIM100 or Cpd4f and probed with anti-ACK1 Tyr(P)284 (PY284) or anti-BRK Tyr(P)342 (PY342) and anti-Flag antibodies. **(D)** May-Grunwald-Giemsa staining of iPSC-derived macrophages from familial controls and ACK1 and BRK patients. Scale bar 10 μm, 100X objective. Representative images from over 50 observed cells per line. **(E) Immunoprecipitation (IP) kinase assay in patients’ macrophages.** (Left panel) ACK1 was immunoprecipitated from BRK^WT/G321R^, ACK1^WT/K161Q^ and ACK1^A156T/K161Q^ iPSC-derived macrophages with anti-ACK1 Ab. The immunoprecipitated proteins were used in duplicate for in vitro kinase reactions with WASP synthetic peptide. Samples of the immunoprecipitates were also probed with anti-ACK1 Ab and anti-tubulin Ab. (Right panel) BRK was immunoprecipitated from ACK1^WT/K161Q^, ACK1^A156T/K161Q^, BRK^WT/G321R^ and BRK^G257A/G321R^ iPSCs-derived macrophages with anti-BRK Ab. The immunoprecipitated proteins were probed with anti-BRK Tyr(P)^342^ (PY342) and anti-BRK antibodies.

ACK1 and BRK sequences are highly conserved in human populations, when compared with other species (**Figure 1C**). Apart from the alleles we report here, only 90 and 35 predicted loss of function (LOF) alleles are reported in gnomAD for ACK1 or BRK respectively, with MAFs ≤ 10^−4^, and none of them reported in homozygosity. Conversely, we examined the *in vitro* kinase activity of ACK1 mutants with a MAF≥0.005 and reported as homozygous in gnomAD (n=4). *In vitro* kinase activity of these variants was normal (**Figure 2-figure supplement 2A-C**). There was no BRK mutant with MAF≥0.005 reported as homozygous in gnomAD. Thus, autosomal recessive deficiency for either protein, whether complete or partial, is expected to be well below 1/100,000 in the general population, which is compatible with autosomal recessive ACK1 and BRK deficiencies underlying SLE in patients from these two kindreds. To investigate whether ACK1/TNK2 or BRK/PTK6 were subject to selection, we gathered data using different metrics quantifying negative selection in the human genome. Analysis of f parameter from SnIPRE ^1^, lofTool ^2^, evoTol ^3^, and CoNeS ^7^ metrics, as well as intraspecies metrics from RVIS ^4^, LOEUF ^5^, and pLI / pRec ^6^ suggested that the genes are not under strong negative selection, consistent with the deficiency being recessive (**Figure 2-figure supplement 2D**).

### ACK1 and BRK inhibition promotes autoimmunity in Balb/c mice

ACK1- and BRK-deficient T cells are characterized by increased proliferation and activation ^36,37^. However, ACK1- and BRK-deficient mice do not develop Lupus-like disease on a C57BL/6 background ^37,38^. We therefore investigated the consequences of inhibiting ACK1 and BRK kinase activity in wild-type (WT) BALB/cByJ female mice which are more susceptible to developing autoimmunity. For this purpose groups of BALB/cByJ mice were either left untreated of received a single intraperitoneal injection of pristane to induce a lupus-like disease ^39^ and a weekly injection of ACK1 or BRK inhibitors Aim100 ^56^ and Cpd4f ^57^, or DMSO vehicle alone for 12 weeks. In the absence of pristane, mice that received ACK1 or BRK inhibitors developed a large array of circulating anti-nuclear IgG antibodies, including but not limited to autoantibodies associated with SLE such as anti-histones, anti-chromatin, anti U1-snRNP, anti-SSA, and anti-Ku (**Fig 3A**). These data suggested that ACK1 and BRK inhibition are sufficient to promote autoimmunity in mice. We did not observe glomerular deposit of IgG at 12 weeks in these mice (**Fig. 3B,C**, **Figure 3-figure supplement 1**). In contrast, BALB/cByJ mice which received pristane treatment ^39^ in addition to either ACK1 or BRK inhibitors had increased kidney glomerular deposits of IgG as well as increased serum autoantibody levels including anti-Ku (p70/p80), LA/SSB, Ro-SSA, Histone H4 and H2B, in comparison to DMSO vehicle controls (**Figure 3A-C**, **Figure 3-figure supplement 1**). Therefore, inhibition of ACK1 or BRK increase serum autoantibody titers and worsen pristane-induced Lupus in WT BALB/cByJ mice. Together with the above genetic analysis, these findings support the hypothesis that autosomal recessive ACK1 and BRK kinase deficiency may underlie or contribute to the development of SLE in children and young adults depending on genetic and environmental context.

**Figure 3.**
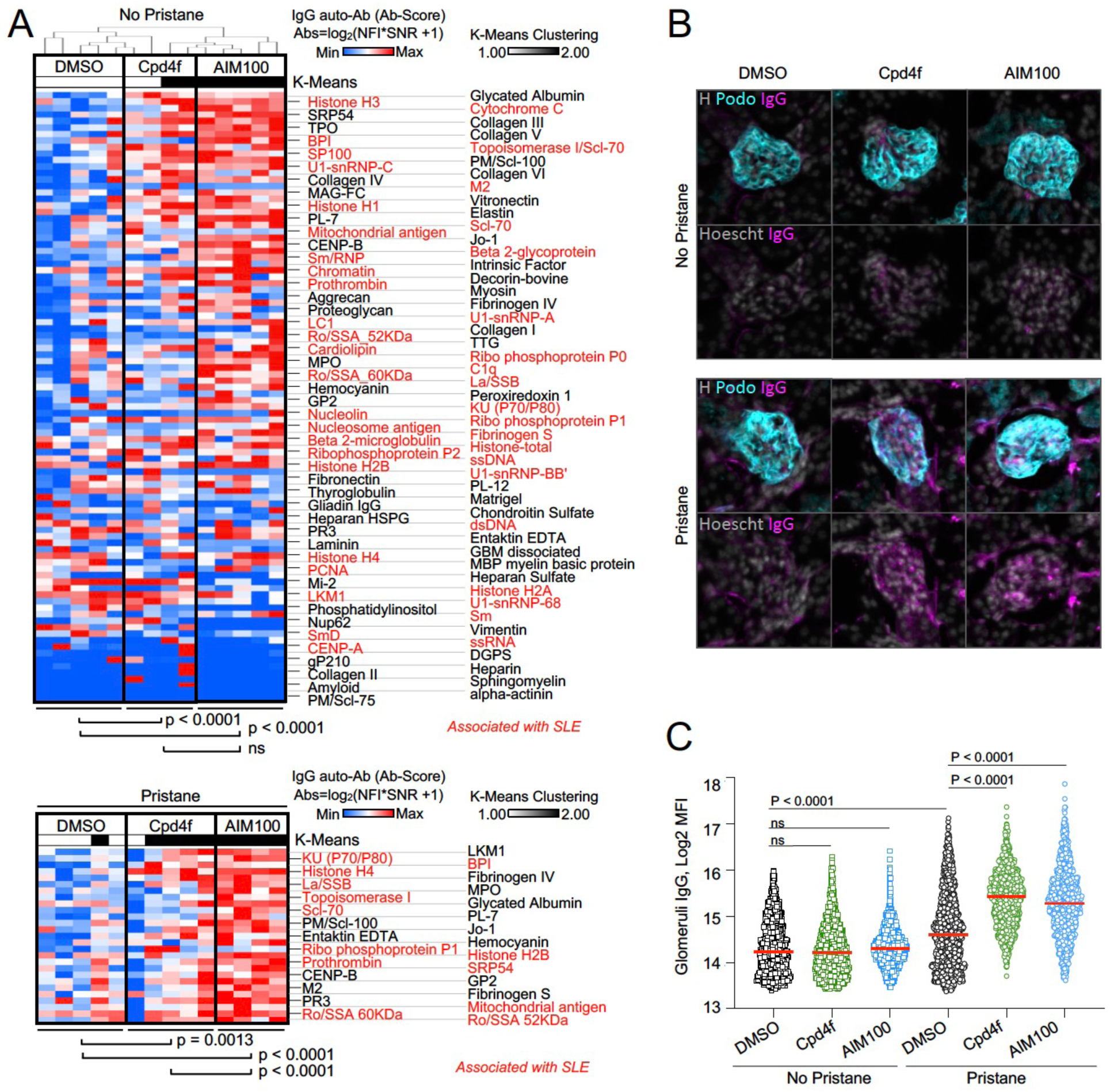
ACK1 and BRK blockade induces autoimmunity in mice. **(A) Heatmaps comparing the levels of IgG autoantibodies detected in serum of mice treated with inhibitors.** Heatmaps show autoantigen microarray panels performed on serum from 4 month-old BALB/cByJ female mice which received a weekly intra-peritoneal injection of DMSO (vehicle, 20ul/mice), AIM100 (25mg/kg in 20ul), or Cpd4f (20mg/kg in 20ul/) since the age of five weeks. *Top panel* depicts results for mice that did not receive a pristane injection. *Bottom panel* represents results for the top differentially produced auto-antibodies in inhibitor treated or control mice that received a Pristane injection at 5 week of age. Plotted values represent Ab Scores (Log_2_ [antigen net fluorescence intensity (NFI) x signal to noise ratio (SNR) +1]. Heatmap columns represent serum analysis of independent mice (n=4-5 for each of the 3 conditions). Heatmap rows sorted top to bottom starting with most significantly increased Ab Score in Cpd4f and AIM100 mice in comparison to DMSO treated mice. P-values were calculated using a Wilcoxon matched-pairs signed rank tests. Hierarchical clustering is based on one minus Pearson correlation with complete linkage method. K-means clustering is based on Euclidean distance, with 2 clusters, with 10000 maximum iterations. **(B,C) Immunofluorescence for mouse IgG on kidney sections.** Representative micrographs (B) displaying glomeruli on kidney sections from 4 month-old BALB/cByJ female mice treated as in (A) and stained with Hoechst 33342, anti-mouse IgG, and anti-mouse Podoplanin antibody. In the quantification plot (C) each symbol represents the IgG mean fluorescence intensity (MFI) in a single glomerulus, of mice treated with designated inhibitors, in the presence or absence of pristane. Approximately 250 glomeruli we analyzed per section/mouse (>95% of all glomeruli in an entire longitudinal kidney section). n=4-5 mice per condition. P-values were obtained using a Kruskal-Wallis test with multiple comparisons.

### ACK1 and BRK kinase domain variants may lose the ability to link MERTK to RAC1, AKT and STAT3 activation for efferocytosis

NRTKs, including ACK1 and BRK, regulate phosphorylation of downstream effectors/ adaptor proteins involved in cell activation, migration, and proliferation including RAC1, AKT, STATs, and ERK ^53,60–66^. NRTK deficiency can result in defective regulation of immune cell activation and survival which can lead to autoimmunity ^36,37,40,67–71^. We found that, in contrast to the reference ACK1 and BRK alleles, the patient’s ACK1 and BRK variant alleles do not phosphorylate AKT and STAT3 (**Figure 4A, B**), and do not activate RAC1 to generate RAC-GTP (**Figure 4C**). NRTKs such as ACK1 ^41^ and PTK2/FAK ^43^ are also downstream targets of the TAM family receptor MERTK which is expressed on macrophages and controls the anti-inflammatory engulfment of apoptotic cells, a process known as efferocytosis ^46,72,73^. Efferocytosis allows for the clearance of apoptotic cells before they undergo necrosis and release intracellular inflammatory molecules, and simultaneously leads to increased production of anti-inflammatory molecules (TGFβ, IL-10, and PGE2) and a decreased secretion of proinflammatory cytokines (TNF-alpha, IL-1β, IL-6) ^46,72–79^. In line with these findings, mice deficient in molecular components used by macrophages to efficiently perform efferocytosis, such as MFG-E8, MERTK, TIM4, and C1q, develop phenotypes associated with autoimmunity ^46,48,72,75–77,79–88^. Furthermore, defects in efferocytosis are also observed in patients with SLE and glomerulonephritis ^75,89–92^

**Figure 4.**
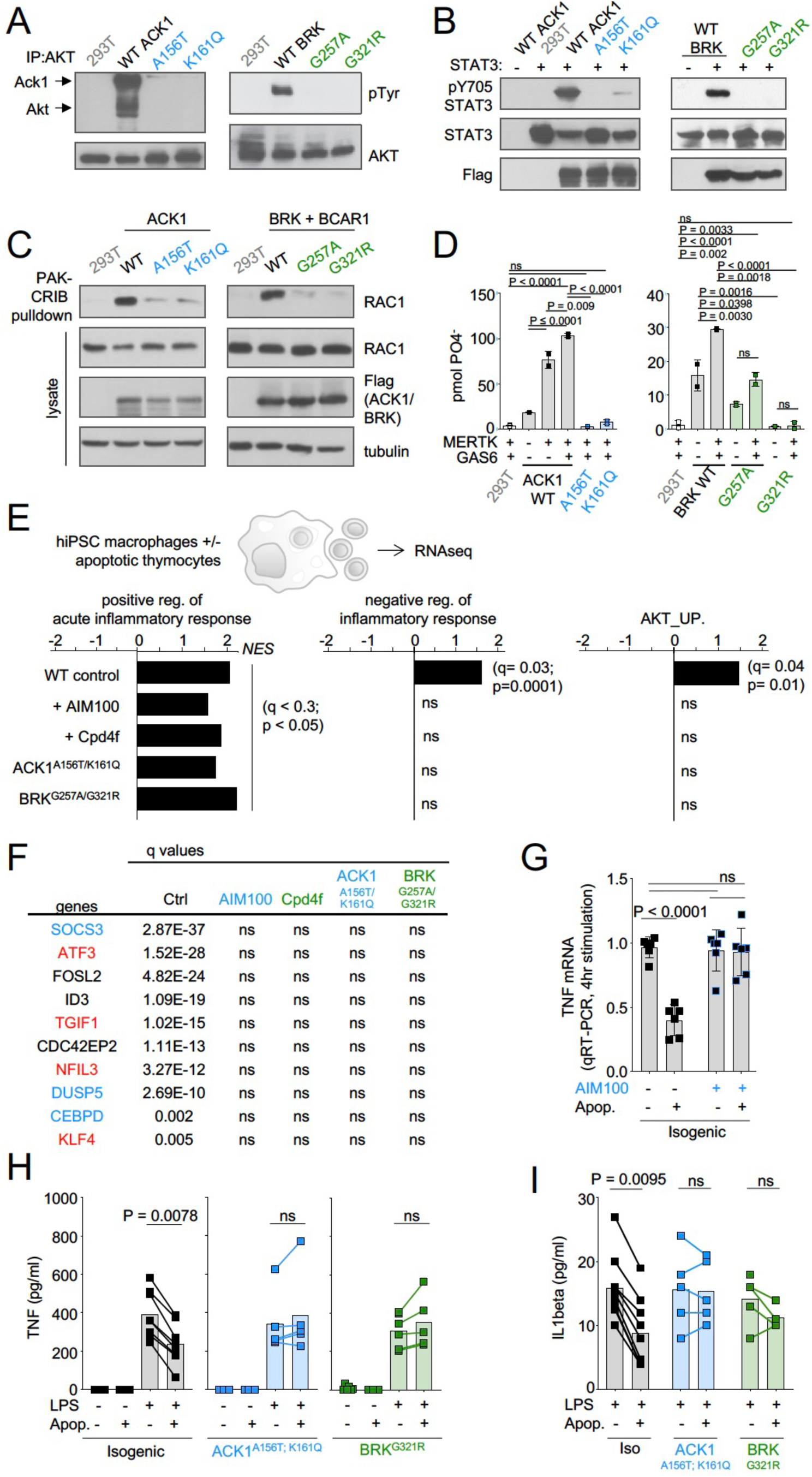
ACK1 and BRK kinase deficiency disrupts the anti-inflammatory response driven by apoptotic cells in macrophages. **(A) Western blot analysis for AKT phosphorylation by ACK1 and BRK.** Cell lysates from 293T cells were incubated with anti-AKT. Immunoprecipitated proteins were probed with anti-phosphotyrosine and anti-AKT antibodies. **(B) Western blot analysis for STAT3 phosphorylation by ACK1 and BRK**. Lysates from 293T cells coexpressing STAT3 and Flag-tagged WT or mutant forms (A156T and K161Q) of ACK1 or mutant forms (G257A and G321R) of BRK were probed with anti-phospho-STAT3 (Tyr705), anti-STAT3 and anti-Flag antibodies. For analysis of BRK, cells were treated with 100 ng/ml EGF for 10 min. **(C) RAC activation by WT ACK1 and BRK.** Cell lysates from 293T cells expressing WT or mutant forms of ACK1 (*left*) and lysates from 293T cells expressing WT or mutant forms of BRK (*right*) were incubated with GST-PAK CRIB sepharose beads, and the level of RAC1 GTP was determined by immunoblotting with anti-Rac1 antibody. Lysates were also probed with anti-Rac1, anti-FLAG and anti-tubulin antibody. For analysis of BRK, 293T cells were cotransfected with CAS and stimulated with 100 ng/ml EGF for 10 min. (**D) MERTK increases kinase activity of BRK and ACK1.** IP kinase assay. ACK1 (left) was immunoprecipitated from 293T cells co-transfected with Flag-tagged ACK1 WT, ACK1 A156T, or ACK1 K161Q and MERTK with anti-Flag Ab. Immunoprecipitated proteins were used in duplicate in vitro for kinase reactions with WASP synthetic peptide and results represented as pmol phosphate transferred. BRK (right) was immunoprecipitated as above from 293T cells co-transfected with Flag-tagged BRK WT or mutants and MERTK with anti-Flag Ab. Kinase reactions was performed with peptide AEEEIYGEFEAKKKG, and represented as above. P-values were calculated using an Anova test (Tukey’s multiple comparison test) (p > 0.05 (ns); p < 0.05 (*); p < 0.01 (**); p < 0.001 (***); p < 0.0001 (****). **(E**) **Regulation of inflammatory response.** Significant normalized enrichment scores (NES) for GO ‘positive regulation of acute inflammation’ gene set, GO ‘negative regulation of inflammatory response’ gene set, and GO ‘AKT_UP.V1_UP’ gene set in WT and mutant macrophages, and WT treated with AIM100 (2 µM) or Cpd4f (0.5 µM), exposed to apoptotic cells, with 3 replicates per experimental condition. Significant enrichment (p-value<0.05 and FDR (q-value) <0.25) are calculated as reported in methods. **(F)** Table of the top 10 differentially regulated genes by apoptotic cells in WT macrophages are not differentially expressed in mutant macrophages and WT macrophages treated with AIM100 or Cpd4f (treated as in E). Numbers indicate FDR (q-value). Known target genes of STAT3 and AKT are labeled in blue and red respectively (**G**) **TNF mRNA production by** WT macrophages treated with AIM100 (2 µM) 4hrs after exposure to apoptotic cells. n=6, from 2 independent experiments. (**H,I) TNF and IL1β production by macrophages**, as measured by ELISA on media collected from mutant and isogenic WT macrophages (C12.1) incubated with mouse apoptotic thymocytes for 90 min, then stimulated with LPS (1 ng/ml) for 18 h. n≥4, from ≥2 independent experiments. P-values in H were calculated by Wilcoxon matched-pairs signed rank tests for data that is not normally distributed, while p-values in G and I were calculated using an *Anova* test with Tukey’s correction for multiple comparisons.

In IP kinase assays MERTK activated the kinase activity of wild-type ACK1 ^41^ but not of the ACK1 A156T and ACK1 K161Q variant alleles (**Figure 4D**). In addition, MERTK also activated BRK kinase activity, but the BRK G321R and G257A alleles were kinase dead and hypomorph variants respectively (**Figure 4D**). MERTK mediates recognition of PtdSer on apoptotic cells via GAS6 and Protein S ^46–48^ leading to their engulfment, which involves activation of RAC1 for actin reorganization and the formation of a phagocytic cup ^43,65^. PtdSer recognition also typically stimulates an anti-inflammatory process mediated in part via AKT ^93^ and STAT3 and their target genes such as SOCS3 ^42,94–99^ and results in the inhibition of LPS-mediated production of inflammatory mediators such as TNF and IL1β, and the production of cytokines such as IL-10, TGFβ ^72,76–79^. Altogether, these data raised the hypothesis that one of the consequences of the defective activity of ACK1 and BRK kinase variants might be an impaired efferocytic response to PtdSer on apoptotic cells.

### ACK1 and BRK kinase deficiency disrupts the anti-inflammatory response driven by apoptotic cells in macrophages

Efferocytosis can be carried out by multiple cell types, however macrophages are the main contributors in this process ^97,100–102^. MERTK kinase activity mediates efferocytosis by human iPSC-derived macrophages ^45^, we thus examined the transcriptional responses of controls, patients, and inhibitor-treated iPSC-derived macrophages to apoptotic thymocytes by RNA-seq (**Figure 4-figure supplement 1**). GSEA analysis of differentially expressed genes indicated that, in contrast to control, ACK1- and BRK-deficient macrophages, as well as WT macrophages treated with ACK1 or BRK inhibitors failed to upregulate gene sets associated with AKT signaling and the negative regulation of the inflammatory response (**Figure 4E**). Transcriptional repressors including the AKT targets ATF3, TGIF1, NFIL3, and KLF4, the STAT3 targets SOCS3 and DUSP5, as well as CEBPD and the inhibitor of E-BOX DNA Binding ID3 were among the top-ten genes which expression is induced by apoptotic cells in WT macrophages (**Figure 4F**), but this regulation was lost in mutant and inhibitor-treated macrophages (**Figure 4F**). ATF3, TGIF1, NFIL3, and KLF4 are involved in the negative regulation of inflammation in macrophages ^94–97^, SOCS3 is an inhibitor of the macrophage inflammatory response and DUSP5 is a negative regulator of ERK activation ^98,99,103^. These data suggest that the kinase domain of ACK1 and BRK contribute to the macrophage anti-inflammatory gene expression program driven by apoptotic cells.

Decreased TNF gene expression by macrophages in response to apoptotic cells was prevented by the ACK1 inhibitor (**Figure 4G**), but the production of TNF at the protein level is not detectable by ELISA in this model (see **Figure 4H**). MERTK-deficient mice are susceptible to LPS-induced endotoxic shock ^104^, and MERTK-dependent anti-inflammatory program elicited by apoptotic cells on macrophages is best evidenced by the reduction of LPS-mediated production of inflammatory mediators such as TNF or IL1β ^76,77,79,93,104^. We thus tested the decrease of LPS-induced production of TNF and IL1β by apoptotic cells. For this purpose, we generated isogenic variants and control hiPSCs and hiPSCs-derived macrophages from the same donor (**Figure 4-figure supplement 2**). Both isogenic variants and control macrophages produced similar amounts of TNF in response to LPS (**Figure 4H**). However, exposure to apoptotic cells inhibited TNF production in isogenic macrophages by 50% but did not inhibit TNF production in ACK1 and BRK-deficient macrophages (**Figure 4H**). Similarly, isogenic variants and control macrophages produced similar amounts of IL1β in response to LPS (**Figure 4I**), but apoptotic cells only decreased IL1β production by isogenic macrophages and not by mutant macrophages (**Figure 4I**). These data altogether indicate that ACK1 and BRK kinase activities contribute to the macrophage anti-inflammatory response during efferocytosis and are required for the decrease of TNF and IL1β production induced by LPS in response to apoptotic cells, a hallmark of their anti-inflammatory effect on macrophages ^72,76,77,79^.

### ACK1 and BRK kinase deficiency alter actin remodeling at the phagocytic cup and modestly decrease engulfment of apoptotic cells in macrophages

MERTK-dependent signaling for anti-inflammatory response and for cargo engulfment driven by recognition of PtdSer on apoptotic cells are distinct and separable molecular events ^44,105^, however because RAC1, which controls engulfment of apoptotic cells ^106^, is a target of ACK1 and BRK (see **Figure 4C**) as well as PTK2/FAK ^43^, we investigated the engulfment of apoptotic cells by ACK1- and BRK-deficient macrophages. We assessed actin-ring formation by total internal reflection fluorescence (TIRF) microscopy in frustrated apoptotic engulfment assays on PtdSer-coated glass slides. Wild-type iPSC-derived macrophages formed a typical actin-ring, however isogenic ACK1 and BRK mutant macrophages presented with an altered actin-ring (**Figure 5A**), and a reduced actin clearance factor (**Figure 5B**), indicating an impairment of actin remodeling following binding to PtdSer. The actin-ring was also altered and actin clearance factor was decreased in WT macrophages treated with ACK1 and BRK inhibitors in comparison to controls (**Figure 5C,D**). These data suggest the kinase activity of ACK1 and BRK contributes to link PtdSer receptors to cytoskeleton rearrangement at the phagocytic synapse.

**Figure 5.**
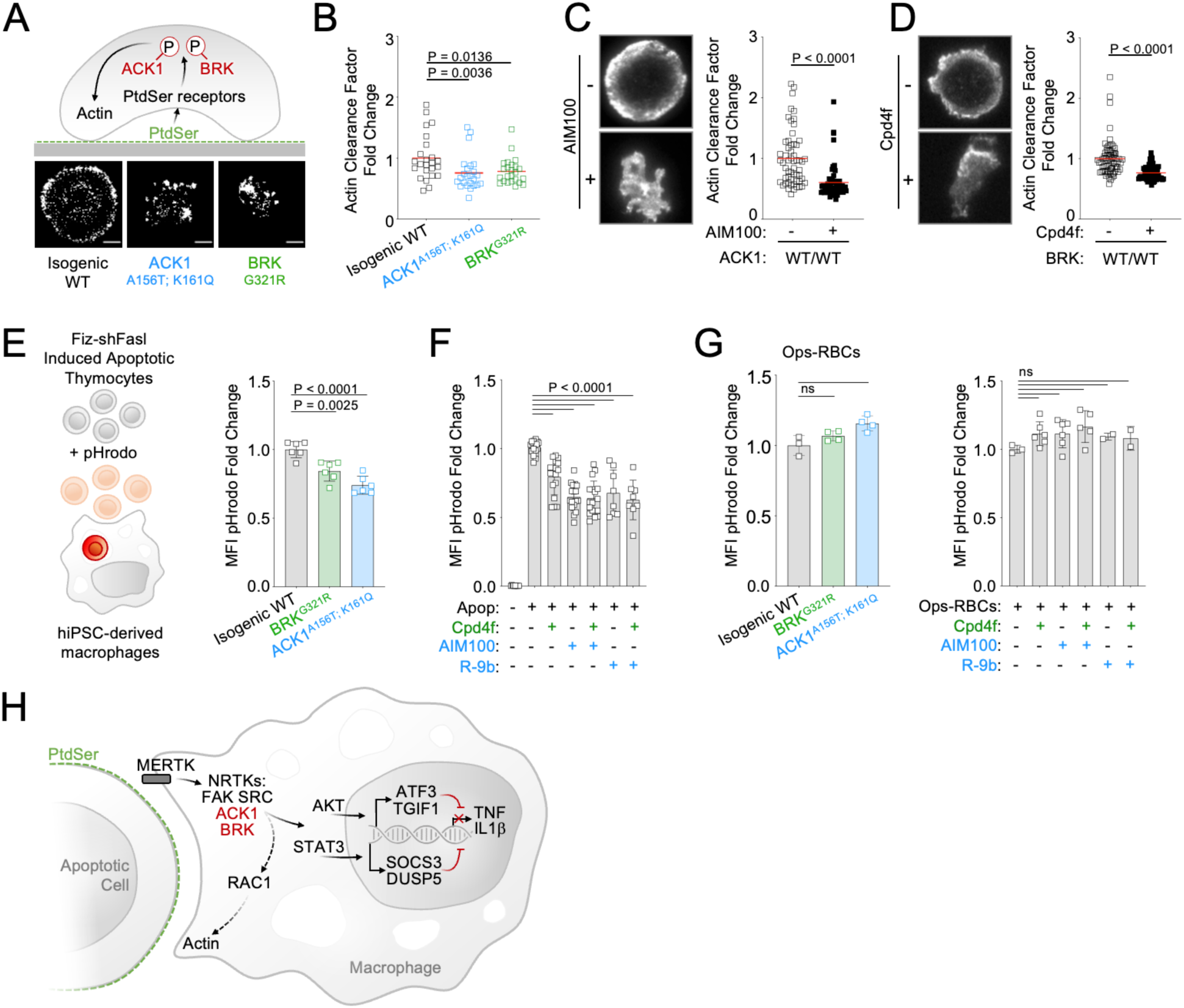
ACK1 and BRK kinase deficiency alter actin remodeling at the phagocytic cup and modestly decrease engulfment of apoptotic cells in macrophages. **(A) Actin remodeling in macrophages.** Schematic and representative images of of F-actin by TIRF microscopy in macrophages of indicated genotype, deposited on PtdSer-coated plates for 20 min. **(B)** Quantification of actin clearance factor for macrophages of the indicated genotypes. Actin remodeling (actin clearance factor) was calculated as a ratio of F-actin staining intensity at cell border divided by F-actin staining intensity at cell center. The actin clearance factor ratios were normalized to the mean value of WT control. Each replicate indicates actin clearance factor fold change from WT mean in single cells. n>20, from 2 independent experiments. Red lines denote the mean. **(C,D) Actin remodeling** quantification (as in A,B) and representative TIRF images of WT macrophages (C12.1 line) pretreated with DMSO, AIM100 (2 µM) or Cpd4f (0.5 µM). n>24, from 3 independent experiments. P-values in B-D were obtained using a Mann-Whitney test. **(E,F) Uptake of apoptotic cells. (E)** Schematic depicts uptake of apoptotic mouse thymocytes treated with Fiz-shFASL and labeled with the pH-sensitive dye pHrodo by iPSC-derived macrophages. Isogenic WT (C12.1 line) and isogenic ACK and BRK point mutant macrophages were incubated with pHrodo-labeled mouse apoptotic thymocytes for 90 minutes and analyzed by flow cytometry. Graph represents mean pHrodo fluorescence Intensity (MFI) fold change calculated by dividing total pHrodo MFI (610/20 nm) of individual samples by the average MFI of isogenic WT macrophages. n≥3, from 3 independent experiments. P-values were obtained using an *Anova* test with Tukey’s correction for multiple comparisons. (**F)** Uptake of apoptotic cells as in (E) with WT macrophages (C12.1 line) pretreated with AIM100 (2 µM), R-9b (4 µM), Cpd4f (0.5 µM), or DMSO. n≥8, from ≥4 independent experiments. **(G) Uptake of opsonized sheep red blood cells.** WT macrophages (C12.1 line) are pretreated as in (F) and incubated with opsonized pHrodo sheep red blood cells for 90 min. Graphs represent mean fluorescence Intensity (MFI) fold change calculated by dividing total pHrodo MFI (610/20 nm) of individual samples by the average MFI of WT macrophages. n≥2, from 2 independent experiments. P-values are obtained using an *Anova* test with Tukey’s correction for multiple comparisons. (**H)** Schematic representation of ACK1 and BRK proposed function in efferocytosis.

We therefore investigated whether this defect translated to a reduced uptake of apoptotic cells by macrophages. We found that engulfment of apoptotic thymocytes labeled with the pH sensitive probe pHrodo ^101,107,108^ was only moderately reduced, by 10 to 20% in ACK1 and BRK isogenic mutant macrophages (**Figure 5E**). This phenotype, although modest, was reproducible by a 30-minute exposure of WT macrophages to two different ACK1 inhibitors, Aim100 (2 µM) and R-9b ^109^ (4 µM), and to the BRK inhibitor Cpd4f (0.5 μM) (**Figure 5F**). This reduced uptake of apoptotic cells was not attributable to a global engulfment defect, because ACK1 and BRK genetic kinase deficiency or inhibitors did not prevent Fc-dependent phagocytosis of a large cargo such as opsonized red blood cells (**Figure 5G**), or the uptake of polystyrene beads (**Figure 5-figure supplement 1A**) or microorganisms such as bacteria and fungi (**Figure 5-figure supplement 1B, C**). Altogether, these data show that the kinase activity of ACK1 and BRK participate to the formation of phagocytic synapse but are largely dispensable for engulfment of apoptotic cells and are not required for phagocytosis of microbes and opsonized cargo.

## DISCUSSION

In this study we combined whole exome sequencing and forward genetic analysis in multiplex SLE families, with a biochemical analysis of genetic variants, murine studies, and functional approaches in human iPSC-derived macrophages to identify novel genes the mutations of which may underlie SLE. In two unrelated families, we identified compound heterozygous loss-of-function or hypomorph variants in the kinase domains of two non-receptors tyrosine kinases, TNK2/ACK1 and PTK6/BRK. Patients from the two families were children or young adults and presented with a severe clinical form of SLE, lupus nephritis. However, we analyzed 27 GWAS studies of SLE (https://www.gwascentral.org/), and none of them reported a common variant in the close vicinity of the TNK2/ACK1 or PTK6/BRK genes with a p-value lower than 5×10^−8^, a statistical threshold of genome-wide significance for GWAS. ACK1 and BRK deficiency are thus likely to only account for the genetic basis of SLE in a minority of patients. Nevertheless, ACK1 and BRK kinase inhibitors aggravate autoimmunity and IgG glomerular deposits in BALB/cByJ mice. Altogether, the present data indicate that autosomal recessive TNK2/ACK1 and PTK6/BRK kinase deficiency probably underlie the development of SLE in a small proportion of children and young adults, depending on genetic and environmental context.

Defective efferocytosis has been shown to contribute to autoimmunity in mice and is relevant to the pathogenesis of SLE ^75,80,81,83–86,89,90^. Our results also suggest that TNK2/ACK1 and PTK6/BRK kinase deficiencies, in addition to dysregulating B and T cell survival and activation ^36,37^, also impairs the MERTK-dependent anti-inflammatory response of the patients’ macrophages to apoptotic cells during efferocytosis, and to a lesser extent the engulfment of the apoptotic cells. MERTK-dependent engulfment of apoptotic cells and anti-inflammatory response were shown to be distinct and separable ^44,105^. The NRTK PTK2/FAK and Src are important for the former ^44,110^, while PTK2/FAK is dispensable for the latter ^44^. Our experiments suggest that in contrast, TNK2/ACK1 and PTK6/BRK are more important for the control of TNF and IL1β production than for engulfment itself. Altogether, our observations identify a rare Mendelian cause of severe SLE and a role for the NRTK TNK2/ACK1 and PTK6/BRK in efferocytosis, thereby contributing to a molecular and cellular dissection of SLE.

## MATERIALS AND METHODS

### Human sample collection and consent information

The study was approved by the Institutional Review Board of St Thomas’ Hospital; Guy’s hospital; the King’s College London University and the Memorial Sloan Kettering Cancer Center. All subject samples were obtained after written informed consent from patients and their families according to the Helsinki convention (Ethics approval: 11/LO/1433). Ten multiplex families with lupus have been enrolled from Guy’s and St Thomas’ NHS Foundation Trust and UCL Hospital in London, UK from July 2010 to January 2012, following stringent criteria: a severe phenotype (lupus nephritis for at least 1 patient in each family), and a familial disease (≥2 family members affected in first degree). A total of 24 patients and 17 healthy controls from different ethnic origins (5 African ancestry, 4 Asian ancestry and 1 European ancestry) were selected. The patients each met the Systemic Lupus International Collaborating Clinics (SLICC) classification criteria for SLE^2^. Lupus nephritis was confirmed by a kidney biopsy classified per the 2004 ISN/RPS (International Society of Nephrology/Renal Pathology Society) classification and verified independently by 2 renal histopathologists.

One hundred Mauritian participants were enrolled under the Ethical Clearance provided by the University of Mauritius Research Ethics Committee. Written consent with due signatures was recorded from all participants prior to partaking in the study. Consent was documented on a confidential form in duplicate, with one copy given to the participants for their records. The University of Mauritius Research Ethics Committee approved, sanctioned and fully endorsed this mode of consent recording. The ethnic backgrounds of the 100 Mauritian participants consisted of 26 Creole, 16 Franco-Mauritian, 21 Indo-Mauritian, 2 Sino-Mauritian, 24 other or undisclosed Mauritians.

### Genetic analysis

Whole Exome Sequencing (WES) of the 10 multiplex families (patients and familial healthy controls) was performed at the New York Genomics Centre on an Illumina HiSeq 2000 sequencing machine. Genomic DNA extracted from the patients and familial healthy control’s peripheral blood cells were sheared with a Covaris S2 Ultrasonicator. An adapter-ligated library was prepared with the Paired-End Sample Prep kit V1 (Illumina). Exome capture was performed with the SureSelect Human All Exon kit (Agilent Technologies). Paired-end sequencing was performed on a HiSeq 2000, generating 100-base reads. For sequence alignment, variant calling and annotation, we used BWA aligner^111^ to align sequences with the human genome reference sequence (hg19 build). Downstream processing was performed with the Genome analysis toolkit^112^, SAMtools^113^, and Picard Tools. Substitution and indel calls were identified with a GATK Unified Genotyper and a GATK Indel GenotyperV2, respectively. All calls with a read coverage ≤2x and a Phred-scaled SNP quality of ≤20 were filtered out. All the variants were annotated with the GATK Genomic Annotator. Variants were annotated following their minor allele frequency (MAF) in Exome Variant Server, 1,000 Genomes Project, and Genome Aggregation Database (gnomAD), including the MAF in each ethnic subpopulation from gnomAD.

First, the variants have been prioritized at the gene level. We used the gene damage index (*GDI*) which provides the accumulated mutational damage of each human gene in healthy human population, based on the 1,000 Genomes Project database (Phase 3) gene variations of healthy individuals and of the CADD score for calculating impact^114^. GDI is very effective to filter out variants harbored in highly damaged (high GDI) genes that are unlikely to be disease-causing. We used a cut-off of 13.84, the recommended GDI value above which a gene is unlikely to be disease-causing which is the 95% CI upper boundary value, removing 5% of the genes in our analysis.

Secondly, we prioritized variants at the allele level. We excluded variants that were too frequent in our in-house database to explain the disease^115^. We then filtered out variants based on the predicted damaging impacts. The deleteriousness of each variant was assessed using in silico algorithms: CADD (http://cadd.gs.washington.edu/score) and Mutation Significance Cutoff browser (MSC, http://pec630.rockefeller.edu/MSC/). MSC scores were generated using 99% confidence interval based on the CADD 1.3 scores of all disease causing-mutations in Human Gene Mutation Database for any given gene^54,55^. We kept only the variants with a CADD/MSC higher than 1.

We thus analyzed our WES with the remaining variants by keeping all non-synonymous coding variants and essential splice site variants with a MAF according to the genetic model: 1) heterozygous variations with MAF <10^−4^ in all ethnic subpopulations under an autosomal dominant (AD) model; 2) homozygous or compound heterozygous variants with MAF <10^−2^ in all ethnic subpopulations) under an autosomal recessive (AR) model; and 3) hemizygous (male) or homozygous (female) variations with MAF <10^−4^ in all ethnic subpopulations under X-linked genetic models.

### Principal component analysis

PCA was performed with 2,504 individuals of the 1,000 Genomes database using WES high quality variants as described in Belkadi et al. We searched for the closest neighbors of the patients in terms of ethnic origin using an Euclidian distance computed from the 10 first PCs ^49,116^.

### Biochemical analysis

#### Cloning and Site-directed mutagenesis

The expression vectors for Flag- and HA-tagged Ack1 and for Flag-tagged Brk were described previously^52,117,118^. Site-directed mutagenesis was performed using the QuikChange Kit (Agilent; 200523). The expression vector for Cas has been described previously^119^. The expression vector for MerTK, pIRES2-EGFP Mer, was a gift from Dr. Raymond Birge^43^.

#### Cell transfection, Immunoprecipitation, and Western Blotting

Cells were transfected 24 h after plating with 8 µL polyethylenimine per µg of DNA in 150 mM NaCl. Cells were harvested 48 h after transfection using lysis buffer (25 mM Tris, pH 7.5, 1 mM EDTA, 100 mM NaCl, 1% NP-40) supplemented with aprotinin, leupeptin, PMSF, and Na_3_VO_4_. For Western blotting, lysates were resolved by SDS-PAGE transferred to PVDF membranes, and probed with the appropriate antibodies. Horseradish peroxidase-conjugated secondary antibodies (GE Healthcare; NA931V; NA9340V) and Western blotting substrate (ThermoFisher Scientific; 32106) were used for detection. Phosphorylation of endogenous Cas is not as strong for BRK as it is for ACK1, so in these experiments we co-transfected with Cas and stimulated BRK by a 10 min 100 ng/ml EGF treatment^53^.

For immunoprecipitation studies, cell lysates (1 mg total protein) were incubated with 1-2 µg of the appropriate antibody and 25 µL of protein A agarose (Roche; 11134515001) for at 4°C for 4 h-overnight. Anti-Flag immunoprecipitations were done with anti-Flag M2 affinity resin (Sigma). The beads were washed three times with lysis buffer, then eluted with SDS-PAGE sample buffer and resolved by SDS-PAGE. The proteins were transferred to PVDF membrane for Western blot analysis.

#### Immunoprecipitation Kinase assay

IP-kinase assays were carried out essentially as previously described^53^. Cell lysates (1 mg protein) were incubated with 25 µL of anti-Flag M2 affinity resin on a rotator for 4°C for 4 h-overnight, then washed three times with Tris-buffered saline (TBS). A portion of each sample was eluted with SDS-PAGE sample buffer and analyzed by anti-Flag Western blotting. The remaining sample was used for a radioactive kinase assay. A WASP-derived peptide (sequence: KVIYDFIEKKKG)^120^ was used as a substrate for Ack1, and a Src-specific peptide (sequence: AEEEEIYGEFEAKKKKG) ^59,121^ was used as a substrate for Brk. The immunoprecipitated proteins were incubated with 25 µL of reaction buffer (30 mM Tris, pH 7.5, 20 mM MgCl_2_, 1 mg/mL BSA, 400 µM ATP), 1 mM peptide, and 50 – 100 cpm/pmol of [γ-^32^P] ATP at 30°C for 20 min. The reactions were terminated using 45 µL of 10% trichloroacetic acid. The samples were centrifuged and 30 µL of the reaction mixture was spotted onto Whatman P81 cellulose phosphate paper. After washing with 0.5% phosphoric acid, incorporation of radioactive phosphate into the peptide was measured by scintillation counting.

#### Rac binding assay

The Cdc42/Rac interactive binding (CRIB) domain from PAK binds specifically to Rac in the GTP-bound state^122^. The PAK-CRIB domain was expressed as a GST fusion protein in E. coli and purified with glutathione-agarose. The immobilized CRIB domain was incubated with cell lysates (1 mg total protein) for 2h at 4°C. The resin was washed with TBS, and bound proteins were eluted with SDS-PAGE sample buffer and analyzed by anti-Rac Western blotting.

### Cells

#### HEK 293T

HEK 293T cells were maintained in Dulbecco’s modified Eagle’s medium (DMEM, Mediatech, Inc.) supplemented with 10% fetal bovine serum (FBS) (Sigma) and 1000 IU/ml penicillin and streptomycin.

#### Derivation of Human iPSCs

Generation of iPSCs from frozen peripheral blood mononuclear cells (PBMCs) was performed using a previously published protocol^126^. Briefly, PBMCs were cultured in QBSF-60 media supplemented with L-Asorbic Acid (50 µg/mL), human SCF (50 ng/mL) (R&D; 255-SC-010/CF), human IL-3 (10 ng/mL) (Peprotech; 200-03), human EPO (2 U/mL) (R&D; 287-TC-500), IGF-1 (40 ng/mL) (R&D; 291-G1-200), and Dexamethasone (1 µM) (Sigma; D8893-1MG) for 9 to 12 days to expand the erythroblast population. Then 4 Sendai viral vectors (ThermoFisher Scientific; A16517) expressing Oct3/4, Sox2, Klf4, or c-Myc are used for transduction of 2.5×10^5^ cells with 10 MOI for each virus for 24 hr. At day 2 post transduction, cells were plated in a 6 well gelatin coated plate containing MEFs (ThermoFisher Scientific; A34181). After 9-12 days, small iPSCs colonies appear. At day 17-21, several colonies were picked and expanded individually into 1 well (12-well or 24-well plate) containing MEFs on gelatin in ESC media as detailed above, supplemented with 10 ng/ml basic fibroblast growth factor (bFGF; Peprotech; 100-18B). Five clones were established per cell line and were maintained in culture for 10 passages (2-3 months) to ensure stability of the lines. Two clones per cell line were selected and tested for chromosomal abnormality and showed a normal karyotype (46, XY or 46, XX). The other clones were frozen down.

#### Culture of Human iPSCs

Human induced Pluripotent Stem Cells (iPSCs) were maintained on mouse embryonic fibroblasts (MEFs, ThermoFisher Scientific; A34181) in ESC media (knock-out Dulbecco’s modified Eagle medium (KO-DMEM, ThermoFisher Scientific; 10829-018) with 20% KO serum replacement (ThermoFisher Scientific; 10828-028), 2 mM L-glutamine (ThermoFisher Scientific; 25030-024), 1% nonessential amino acids (ThermoFisher Scientific; 11140-035), 1% penicillin/streptomycin (ThermoFisher Scientific; 15140-122), 0.2% β-mercaptoethanol (ThermoFisher Scientific; 31350-010) supplemented with 10 ng/ml basic fibroblast growth factor (bFGF, Peprotech; 100-18B). Passaging was performed every 7 days at 1:3-1:6 dilution ratio depending on the colony size. During passaging, iPSCs are detached as clusters by a 13 min incubation at 37°C with collagenase type IV (250 UI/ml final concentration) (ThermoFisher Scientific; 17104019) and are pelleted at room temperature by centrifugation at 100G. The iPSC clusters are resuspended in ESC medium supplemented with 10 ng/ml bFGF (Peprotech; 100-18B) and plated on NUNC plates containing 12,500 to 16,000 MEFs per cm^2^.

#### Generation of the TNK2 (ACK1) and PTK6 (BRK) Isogenic Mutant iPSC lines

CRIPSR and single-stranded donor oligonucleotides (ssODN) were used as the tool to introduce the SNP mutations in hiPSCs. CRISPR sgRNA target was designed using the web resource at https://www.benchling.com/crispr/. The target sequence was cloned into the pX330-U6-Chimeric_BB-CBh-hSpCas9 vector (Addgene plasmitd #42230) to make the gene targeting construct. The template ssODN was designed to carry the mutant nucleotide and served as the donor template. The ssODN was then purchased from IDT. The sgRNA-target and ssODN sequence are listed in Table 2.

**Table 2:**
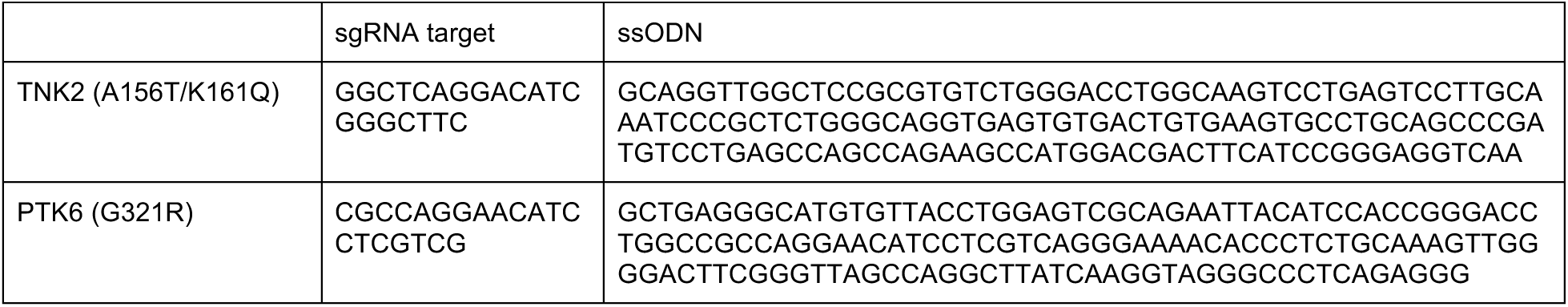

To introduce the SNP mutations, WT iPSCs (C12) were dissociated using Accutase (Innovative Cell Technologies) and electroporated (1×10^6^ cells per reaction) with 4 µg sgRNA-construct plasmid and 4 µl ssODN (10 µM stock) using Human Stem Cell Nucleofector^TM^ solution (Lonza) following manufacturer’s instructions. The cells were then seeded, and 4 days later, hESCs were dissociated into single cells by Accutase and re-plated at a low density (4 per well in 96-well plates) to get the single-cell clones. 10 days later, individual colonies were picked, expanded and analyzed by PCR and DNA sequencing. The PCR and sequencing primers are listed in Table 3.

**Table 3:**
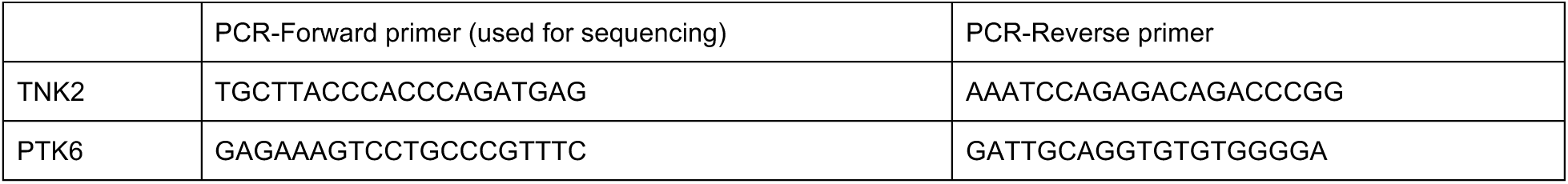

#### Differentiation of iPSCs-derived macrophages

The hiPSCs to macrophage differentiation method was adapted from a previously published protocol^58^. Briefly, newly passaged iPSCs were maintained from day 0 to day 3 in ESC media (see human iPSCs culture) with 10 ng/ml bFGF and from day 3 to day 7 in ESC media without bFGF. At day 7, iPSCs colonies were detached in clusters using collagenase type IV (250 UI/ml final concentration) (ThermoFisher Scientific; 17104019) and transferred to 6 well suspension plates in ESC media supplemented with 10 μM ROCK Inhibitor (Sigma; Y0503), on an orbital shaker at 100 rpm. The cell clusters were cultivated under these conditions from day 7 to day 13 to induce embryoid body (EB) formation. At day 13, well formed 200-500 μm EBs were manually picked under a microscope and transferred onto adherent tissue culture plates (~2.5 EBs/cm^2^) for cultivation in APEL 2 medium (Stem Cell Tech; 05270) supplemented with 5% protein free hybridoma (ThermoFisher Scientific; 12040077), 100 IU/ml penicillin and 100 μg/ml streptomycin (ThermoFisher Scientific; 15140-122), 25 ng/ml human IL-3 (Peprotech; 200-03) and 50 ng/ml human M-CSF (Peprotech, 300-25). Starting from day 25 of the differentiation and then every week onwards for up to 5 weeks, suspension cells around EBs were carefully collected, filtered through a 100 µm mesh, plated at a density of ~15000 cells/cm^2^, and cultivated for 6-10 days in RPMI1640/GlutaMax (ThermoFisher Scientific; 61870036) medium supplemented with 10% FBS (EMD Milipore TMS-013-B), and 100 ng/ml human M-CSF and used further for functional analysis. All cells were cultured at 37°C 5% CO_2_ in standard tissue culture incubators.

### Mice

C57BL/6J mice were used for preparation of apoptotic thymocytes and BALB/cByJ female mice were used for *in vivo* inhibitor treatment experiments (see below). Mice were purchased from The Jackson Laboratory. All mouse studies were performed in adherence with Institutional Review Board (IACUC 15-04-006 and 13-04-003) from MSKCC.

### Antibodies and Flow cytometry

The following antibodies were used for human iPSCs-derived macrophages phenotyping: PE/Cy7 anti-human MERTK Ab (Biolegend; 367609); PE/Cy7 anti-human CD11b Ab (BioLegend; 301321); PE anti-human Intα5β3 Ab (R&D; FAB3050P); APC anti-human TIM4 Ab (BioLegend; 354007); AF647 anti-human TIM4 Ab (BioLegend; 354007); APC/Cy7 anti-human CD36 Ab (Biolegend; 336213); BV786 anti-human CD115 Ab (CSF-1R) (BD Biosciences; 743145); PE-Cy5 anti-human CD11c Ab (BD Biosciences; 561692); Alexa Fluor 700 anti-human HLA-DR Ab (BD Biosciences; 560743); APC/Cy7 anti-human CD45 Ab (BioLegend; 304014); BV650 anti-human CD14 Ab (BioLegend; 301836); Alexa Fluor 647 anti-human CD369 (Clec7A) Ab (BD Biosciences; 564855); Alexa Fluor 488 anti-human CD206 (MRC1) Ab (ThermoFisher Scientific; 564855)

iPSCs-derived macrophages were detached using trypsin (TrypLE Express, ThermoFisher Scientific; 12605-010, pelleted at 400g for 5 min and resuspended in fluorescence-activated cell sorting (FACS) buffer (PBS +0.5% bovine serum albumin (BSA) +1 mM EDTA). After blocking Fc receptors (Miltenyi; 130-059-901) at 1/10 dilution for 10 min, the cells were washed with FACS buffer, pelleted at 400G for 5 min, and immunostained in FACS buffer + antibody (1:50 to 1:200 dilution) for 30 min at 4°C. Data were acquired on an ARIA III BD flow cytometer or a FACS Fortessa SORP instrument and analyzed with FlowJo. Dead cells and debris were excluded from the analysis using DAPI (1 µg/ml), side (SSC-A) and forward scatter (FSC-A) gating, and doublet exclusion using forward scatter width (FSC-W) against FSC-A. At least 5000 cells were acquired for each condition.

### Cytology

Cells were collected into FBS and centrifuged (800 rpm, 8 min, low acceleration) onto Superfrost slides (ThermoFisher Scientific) using a Cytospin 3 (Thermo Shandon). Slides were air-dried for at least 30 min, and fixed for 5 min in methanol, stained in 50 % May-Grunwald solution for 15 min, 5% Giemsa for 15 min, washed with Sorensons buffered distilled water (pH 6.8) for 5 min and rinsed with Sorensons buffered distilled water (pH 6.8). Slides were air-dried and mounted with Entellan New (Merck) and representative pictures were taken using an Axio Lab.A1 microscope (Zeiss) under a N-Achroplan 100x/01.25 objective.

### Engulfment of Beads, E. coli, or C. albicans by iPSC-macrophages

WT iPSCs-derived macrophages were plated at a density of 35,000 cells per well in 24 well plates and maintained in 0.5 ml of RPMI containing 10% FBS. The macrophages were pretreated for 30 min with AIM100 (2 μM) and/or Cpd4f (0.5 μM) in RPMI without FBS prior to incubation with beads, E.coli or C. albicans.

Red fluorescent 2 μM Beads (Microparticles) (Invitrogen, F8826) were resuspended in RPMI (5×10^5^ beads/ml), sonicated for 5 min, and part of the suspension was warmed to 37°C or cooled to 4°C. 400 μl of the suspension was added per well (2×10^5^ beads/well) to pretreated iPSC-macrophages for each condition, and the plates were incubated for 1 h at 37°C or 4°C.

pHrodo E. coli BioParticles (Invitrogen, P35361) were resuspended in RPMI (0.5 mg/ml), sonicated for 5 min, and part of the suspension was warmed to 37°C or cooled to 4°C. 400 μl of the suspension was added per well (0.2 mg/well) to pretreated iPSC-macrophages for each condition, and the plates were incubated for 1 h at 37°C or 4°C.

Nonfluorescent and tdTomato positive C. albicans (clinical isolate SC5314) were grown overnight in YPD media at 30°C with 225 rpm shaking. The cells were washed with PBS, spun at 1000G for 5 min, and resuspended in RPMI to a density of 740,000 cells/ml. The suspensions were then warmed to 37°C or cooled to 4°C. 500 µl of the suspension was added per well (370,000 C. albicans cells) of iPSC-macrophages for each condition, and the plates were incubated for 1 h at 37°C or 4°C.

After the incubation with beads, E. coli or C. albicans, the media was removed and the wells were washed with PBS. The cells were then detached with a 3 min, 37°C incubation with trypsin, and collected by centrifugation at 400G for 5 min. The iPSC-macrophages incubated with beads or E. coli were resuspended in 200 μl of FACS buffer. The cells incubated with C. albicans were resuspended in PBS containing 20 μg/ml Calcofluor White (CFW) stain (Sigma), and were incubated for 15 min at room temperature to label unengulfed yeast. The cells were then washed with FACS buffer and pelleted at 400G for 5 min at 4°C, before being resuspended in 200 μl of FACS buffer.

The samples were analyzed using a FACS Fortessa SORP instrument. pHrodo Red and tdTomato fluorescence was detected through a 586/15 bandpass optical filter, on a 561nm laser excitation. CFW was excited by a 405 nm laser, and detected through a 495LP, 525/50 bandpass optical filters. At least two replicates were done for each condition.

### Engulfment of murine apoptotic thymocytes and opsonized red blood cells

Engulfment of apoptotic cells was assayed with pHrodo-labeled mouse apoptotic thymocytes as previously described^107,127^. Briefly, thymocytes from 4- to 8-week-old C57BL/6J mice were treated with Human leucine-zipper-tagged Fas ligand (FIZ-shFasL) (see below for preparation and concentration) in RPMI1640 containing 10% FBS for 2 h at 37°C to induce apoptosis, washed with PBS, and incubated with 0.1 μg/ml pHrodo for 30 min at room temperature. After the reaction was stopped with 1 ml FBS, the cells were washed with PBS containing 10% FBS and were used as prey.

Sheep red blood cells (MP Biomedicals, 0855876) were washed two times in PBS at 600G for 5 min, at room temperature. The cells were resuspended in PBS, counted and further diluted to a concentration of 1×10^8^ cells/ml. To opsonize the RBCs, 1/500 dilution of Rabbit Anti-Sheep RBC IgG (Cell Biolabs; 122001; CBA-220) was added and the cells were incubated at 37°C for 30 min. The cells were then washed twice with PBS at 600G for 5 min, and stained in 1ml of PBS with 10 µg/ml pHrodo, under a 45 min incubation at room temperature. 1 ml of FBS was added to block the staining, and the cells were washed with PBS containing 10% FBS before being used in engulfment.

0.5×10^6^ pHrodo-labeled apoptotic cells, or opsonized RBCs were added to 60 000 iPSCs-derived macrophages in 0.75 ml of RPMI containing 10% FBS in a 12-wells plate, and incubated at 37°C for 90 min. The cells were washed with PBS and detached with trypsin (TrypLE Express, ThermoFisher Scientific; 12605-010). The cells were collected by centrifugation at 400 g for 5 min, suspended in 300 μl of CHES (N-cyclohexyl-2-aminoethane-sulfonic acid)–fluorescence-activated cell sorter (FACS) buffer (20 mM CHES buffer [pH 9.0] containing 150 mM NaCl and 2% FBS) and analyzed by flow cytometry with a FACS Fortessa SORP instrument. At least 5000 cells were acquired for each condition. pHrodo was excited with the yellow green laser 561 nm and detected with 610/620 nm bandpass filter.

FIZ-shFasL was produced using HEK293T as described previously^108^ and concentrated (around 50 fold) by ultrafiltration with Amicon Ultra-15 10K column (Sigma; UFC901008). The concentration used from each batch was determined with Alexa Fluor 488 Annexin V/Dead Cell Apoptosis Kit, as the concentration for which more than 80% of cells were annexin V positive and less than 10% propidium iodide positive.

### Frustrated phagocytosis assay and TIRF imaging

Supported lipid bilayers containing 1,2-dioleoyl-sn-glycero-3-phospho-L-serine (DOPS; Avanti Polar Lipids, 840035) were prepared as previously described ^128^. Cells were incubated on bilayers (20 min, 37°C) and fixed by adding 3% paraformaldehyde (20 min). Fixed cells were permeabilized with 0.2% Triton X-100 (15 min) and blocked in 10% goat serum/PBS (1 hr). Then cells were incubated with the phospho-Cas (Tyr165) antibody (Cell Signaling; 4015) (16 h, 4°C), washed, and incubated with the Alexa Fluor 488 goat anti-rabbit secondary antibody and 0.1 U/ml Alexa Fluor 594–labeled phalloidin (ThermoFisher Scientific; A12381) (1 hr, room temperature). TIRF images of fluorescently labeled cells in contact with bilayers were collected with a 60× objective lens (1.45 NA; Olympus) using 488 and 561 nm lasers (Melles Griot) for imaging of Phospho-CAS (Cell signaling; Cat#4015) and Phalloidin (ThermoFisher Scientific; Cat#A12381), respectively.

Phospho-CAS intensity was calculated as the background-corrected mean fluorescence intensity (MFI) in every cell area, manually drawn using SlideBook software (3I). Quantification of clearance ratio was performed with Matlab software (Mathworks) from the TIRF images of background-corrected MFI as previously described^129^. Briefly, using two perpendicular linescans for each cell, the background-corrected MFI at the edges (positions F1 and F2) of the IS was compared with the background-corrected MFI of three equally spaced central positions (F3, F4, and F5) as follows: mean (F3 + F4 + F5) / mean (F1 + F2). Clearance ratios derived from the two-perpendicular line-scans were averaged to yield a clearance ratio for the cell in question.

### Transcriptomic analysis by RNA-seq

70,000 iPSCs-derived macrophages were plated per well in 12 well plates in RPMI1640/GlutaMax medium supplemented with 10% FBS, and 100 ng/ml human M-CSF. WT and mutants (ACK1A156T/K161Q and BRKG257A/G321R) iPSCs-derived macrophages were co-incubated or not with 450,000 mouse apoptotic thymocytes (as described above) in RPMI for 90 min. WT iPScs-derived macrophages were also pretreated for 30 min with AIM100 (2 μM) or cpd4f (0.5 μM) in RPMI and then incubated or not with mouse apoptotic thymocytes. Every condition was done in duplicate or triplicate. After 90 min incubation media was removed and macrophages were washed once with PBS. Then 1 ml TRIzol (ThermoFisher Scientific; 15596018) was added per well and cells were harvested and stored at −80°C. RNA from cells suspended in TRIzol was extracted with chloroform. Isopropanol and linear acrylamide were added, and the RNA was precipitated with 75% ethanol. Sample were resuspended in RNase-free water.

#### Transcriptome sequencing

After RiboGreen quantification and quality control by Agilent Bioanalyzer, 65.8-100 ng of total RNA underwent polyA selection and TruSeq library preparation according to instructions provided by Illumina (TruSeq Stranded mRNA LT Kit; RS-122-2102), with 8 cycles of PCR. Samples were barcoded and run on a HiSeq 4000 or HiSeq 2500 in rapid mode in a 50bp/50bp paired end run, using the HiSeq 3000/4000 SBS Kit or HiSeq Rapid SBS Kit v2 (Illumina). An average of 50 million paired reads was generated per sample. At the most the ribosomal reads represented 7.1% of the total reads generated and the percent of mRNA bases averaged 80.4%.

#### Analysis

The output data (FASTQ files, see below) were mapped to the target genome using the rnaStar aligner^130^ that both maps reads to the genome and resolves reads that map across splice junctions. We used the 2 pass mapping method outlined as described^131^, in which the reads are mapped twice. The first pass used a list of known annotated junctions from Ensemble. Novel junctions found in the first pass are then added to the list of known junctions and then a second mapping pass is done (on the second pass the RemoveNoncanoncial flag is used). After mapping we post process the output SAM files using the PICARD tools to: add read groups, AddOrReplaceReadGroups which in addition sorts the mapped reads by coordinates and coverts the file to the compressed BAM format. We then compute the expression count matrix from the mapped reads using HTSeq (www-huber.embl.de/users/anders/HTSeq) using Genecode v18 database for gene models. The raw count matrix generated by HTSeq was then processed using the R/Bioconductor package DESeq (www-huber.embl.de/users/anders/DESeq) which was used to both normalize the full dataset and analyze differential expression between sample groups.

The hypergeometric test and Gene Set Enrichment Analysis (GSEA)^132^ was used to identify enriched signatures using the different pathways collection in the MSigDB database^133^. We used GSEA pre-ranked method from GSEA for our purpose.

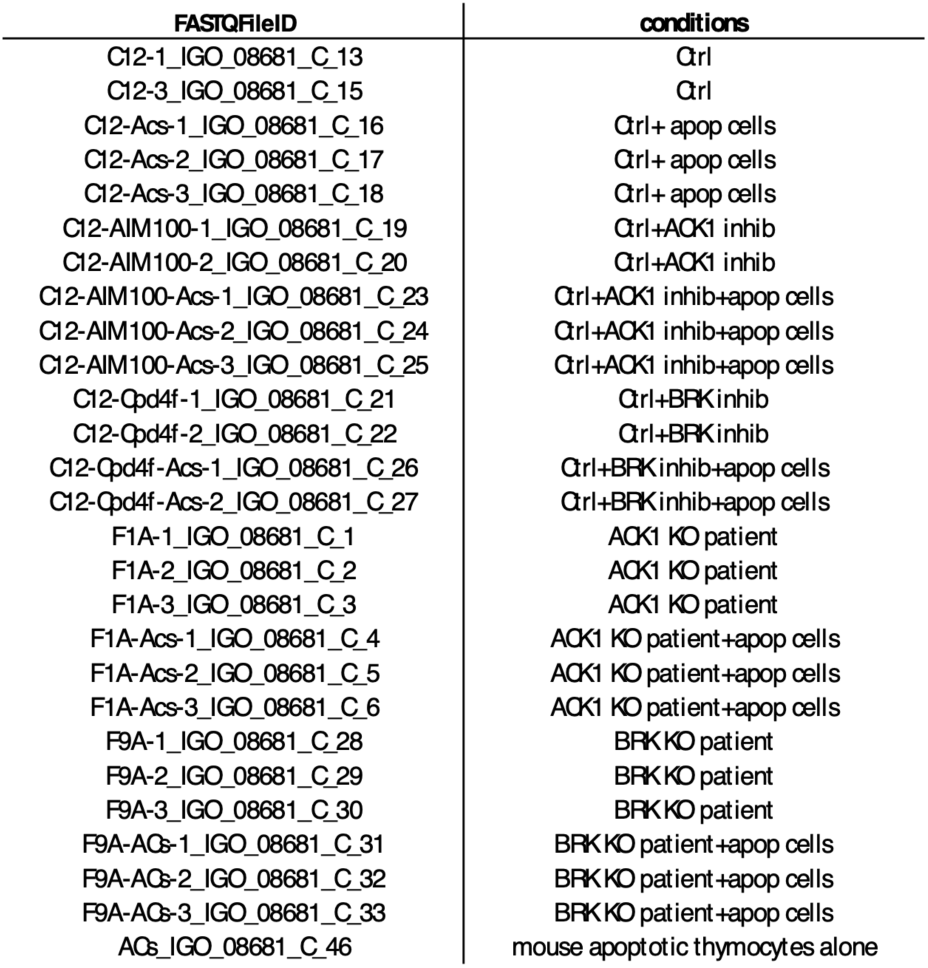

### Quantitative RT-PCR

Total RNA was extracted from cells using a quick-RNA Microprep kit (Zymo research; R1050) as per manufacturer’s instructions. RNA was extracted from 150,000 iPSCs-derived macrophages in 6 well plates or 37500 iPSCs-derived macrophages in 24 well plates depending on the quantity of RNA needed. Lysis buffer was added directly in the well after 1 wash with PBS, then RNA was extracted directly or cells in lysis buffer were stored at −80°C. cDNA preparation was performed with Quantitect Reverse transcription kit (Qiagen; 205313) as per manufacturer instructions. qRT-PCR are done with 20 ng cDNA. qRT-PCR are performed on a Quant Studio 6 Flex using TaqMan Fast Advance Mastermix (ThermoFisher Scientific; 4444557), and TaqMan probes for GAPDH (Hs02758991_m1), TIMD4 (Hs00293316_m1), MERTK (Hs01031973_m1), ITGB5 (Hs00174435_m1), ITGB1 (Hs01127536_m1), ITGB3 (Hs01001469_m1), TNK2 (Hs01006880_m1), PTK6 (Hs00966641_m1), TNF (Hs00174128_m1). Comparative threshold cycles (CT) was used to determine gene expression. For each sample, genes CT value were normalized with the formula ΔCT = CTgene − CTGAPDH. For relative expression, the mean ΔCT was determined, and relative gene expression was calculated with the formula 2^(−ΔCT).

### TNF and IL1β ELISA

hiPSC macrophages plated in 24 well tissue culture plates (density of 15000 cells/cm2) were co-cultured with or without apoptotic thymocytes (1/8 mac./apop.t. ratio) for 90 min at 37C in complete media (RPMI, 10% FBS, 100ng/ml M-CSF). The wells were then washed once with PBS at 37°C to remove the majority of remaining apoptotic thymocytes. 300ul of complete media with or without 1ng/ml LPS was added in each well and the cells were incubated for 18hrs at 37C in a CO_2_ incubator. The media was then collected and stored at −80C. ELISA was conducted according to manufacturer’s protocol (Human TNF-alpha DuoSet (R&D; DY210) and IL1β DuoSet (R&D, DY201) ELISA kit).

### *In Vivo* Mouse Inhibitor and Pristane Treatment, Serum Preparation, and Kidney Fixation

Five week-old BALB/cByJ female mice received intra-peritoneal injection of DMSO (vehicle, 20 µl/mice), AIM100 (25 mg/kg in 20 µl), or Cpd4f (20 mg/kg in 20 µl) for 3 months. Injection were administered biweekly for the first 5 weeks, and weekly subsequently. The mice received a single intraperitoneal injection of PBS (500ul) or pristane oil (Sigma, P9622) (500ul) when 7 weeks old. Blood was collected by cardiac puncture. To collect serum, the blood was left to clot, undisturbed at room temperature for 30 min. The samples were then centrifuged at 1500G for 10 min at 4°C and the supernatant/serum was collected and stored at −80°C. The kidneys were removed, washed with PBS at 4°C, and were fixed whole in 4% PFA overnight at 4°C. The kidneys were then washed with PBS and place in 15% sucrose at 4°C for 3.5hrs. The kidneys were transferred in 30% sucrose and incubated overnight at 4°C. The kidneys were then dried with kimwipes and embedded in blocks in frozen section compound (Leica Ref:3801480). The blocks were placed in dry ice ethanol bath to freeze and were transferred to −80°C for storage.

### Kidney Histology and Image Analysis

Frozen kidneys were sectioned longitudinally into 12um sections and transferred on glass slides. The sections were left to dry for 20-30minutes at room temperature (RT) and were transferred for storage at −20°C. To stain, the sections were thawed at RT for 15 minutes in a humidified box, and rehydrated by washing in PBS at RT 3 times. The sections were incubated for 1hr at RT in PBS, 5% BSA to block. The sections were then washed 2 times with PBS at RT and stained overnight at 4°C with IgG AF555 (Goat Anti-Mouse; ThermoFisher A21424) (1/500 dilution in PBS, 0.5% BSA), and Podoplanin (Syrian Hamster Anti-Mouse; Biolegend 127401) (1/100 dilution in PBS, 0.5% BSA). The sections were washed with PBS at RT 3 times and were incubated for 1.5hrs at RT with Anti-Syrian Hamster AF647 (Jackson, 107-605-142) (1-500 in PBS, 0.5% BSA). The sections were washed with PBS at RT 3 times and stain with Hoechst (ThermoFisher 62249) (20uM final concentration in PBS) for 10 min at room temperature. The slides were washed with PBS 2 times, and mounted using 170ul of ProLong Gold antifade reagent (Invitrogen P36930) and high precision microscope cover glass (24X50mm; 170uM; No. 1.5H). Entire kidneys sections were imaged on confocal microscope with 10X objective.

To analyze IgG mean fluorescence intensity (MFI) of glomeruli, podoplanin staining was used to algorithmically generate outlines around glomeruli in full kidney section images using ImageJ. The outlines were manually verified to remove incorrectly marked glomeruli and to outline the glomeruli the algorithm missed. Over 95% of all the glomeruli in a section (about 250 per whole longitudinal kidney section) were captured and analyzed for each mice (5 mice were analyzed for each condition (30 whole kidney sections in total)). The MFI of each outlined glomeruli was extracted with ImageJ.

### Autoantigen Microarray Panel Profiling

For each sample, 10 µl of serum was treated with DNAse I, diluted 1:50, and incubated with autoantigen array. The autoantibodies binding to the antigens on the array were detected with Cy3 labeled anti-IgG. The arrays were scanned with GenePix® 4400A Microarray Scanner and the images were analyzed using GenePix 7.0 software to generate GPR files. The averaged net fluorescent intensity (NFI) of each autoantigen was normalized to internal (IgG) controls. Plotted values represent Ab Scores (Log_2_ [antigen net fluorescence intensity (NFI) x signal to noise ratio (SNR)+1]. Heatmaps were plotted with Morpheus (https://software.broadinstitute.org/morpheus/). Heatmap rows were sorted top to bottom starting with most significantly increased Ab Score in Cpd4f and AIM100 in comparison to DMSO treated mice. Single row P value between DMSO and Cpd4f or AIM100 mice was calculated by paired t test. Full panel P-values were calculated using a paired t test. Hierarchical clustering was done by one minus Pearson correlation with complete linkage method. K Means clustering was done by Euclidean distance, with 2 clusters, with 10000 maximum iterations.

### Quantification and Statistical Analysis

Data are shown as mean with individual values or as bar plots with mean values and standard deviations. Statistical significance was analyzed with GraphPad Prism unless otherwise indicated (see below). Statistical significance was determined using a Student t test, Mann-Whitney, ordinary one-way ANOVA with Tukey’s multiple comparisons test, or Kruskal-Wallis multiple comparisons tests depending on the number of conditions being compared and whether the data was normally distributed as determined by a Shapiro-Wilk normality test. P < 0.05 values were considered statistically significant. R software was used for statistical analysis of RNA-seq data. For differential gene expression approach significance was considered for FDR (q value) < 0.05. For gene set enrichment analysis (GSEA) approach significance was considered for FDR (q value) < 0.25 AND p-value < 0.05.

## Data and materials availability

Genealogy trees of the 10 multiplex families recruited, Clinical features of the 22 lupus patients recruited from 10 kindreds, and Clinical features of the patients in ACK1 and BRK families can be requested from the corresponding author.

Raw data files for the RNA sequencing analysis have been deposited in the NCBI Gene Expression Omnibus under accession number GEO: GSE118730.

## Contact for reagent and resource sharing

Further information and request for reagents may be directed to and will be fulfilled by the corresponding author Frederic Geissmann.

## Data Availability

Raw data files for the RNA sequencing analysis have been deposited in the NCBI Gene Expression Omnibus under accession number GEO: GSE118730. Further information and requests may be directed to and will be fulfilled by the corresponding author Frederic Geissmann.

## ACKNOWLEDGEMENTS

**General:** We are grateful to Ingeborg Bajema and Suzanne Wilhelmus from Leiden University Medical Center and Terry Cook from Imperial College Healthcare NHS trust London for help with the pathological review of patients, and Louise Nel from Guy’s and St Thomas’ NHS Foundation Trust for taking care of the patients.

## Funding

We acknowledge the use of the MSKCC Stem Cell Research Core and MSKCC Integrated Genomics Operation Core, funded by the NCI Cancer Center Support Grant (CCSG, P30 CA08748), Cycle for Survival and the Marie-Josée and Henry R. Kravis Center for Molecular Oncology. This work was supported by National Cancer Institute of the US National Institutes of Health (P30CA008748) MSKCC core grant and grants from Ludwig Institute for Cancer Research and NIH/NIAID 1R01AI130345-01 and 5R01AI124349-03 (to FG) NIH/NIAID R01-AI087644 (to MH), NIH/NCI CA58530 (to WTM), NIH/NIAID R01-AI087644 to MH, and Grants-in-Aid for Scientific Research (S) from JSPS (No. 15H05785) and Core Research for Evolutional Science and Technology from Japan Science and Technology Agency (JPMJCR14M4) (to SN). SG was supported by Fellowships from the Fondation pour la Recherche Medicale (DEA20140630127), the European Federation of Internal medicine (EFIM), the Assistance Publique-Hopitaux de Paris (Annee Recherche), and from Institut Servier. NJ was supported by Fellowships from the Arthritis Research UK Fellowship and Graham Hughes Clinical Research Fellowship; her present address: Rheumatology Department, Cambridge University Hospitals, Hills Road, Cambridge, CB2 0QQ, UK.

## Author contributions

FG designed the study. SG, TL and FG wrote the manuscript and prepared figures. NJ, DDC and DI selected patients and collected DNA and cell samples. SG, BB, LA, and JLC performed and analyzed exome sequencing. SDD and RIFL collected and performed exome sequencing of Mauritian participants. NL provided background in iPSC generation and macrophage differentiation. WTM and BC performed and analyzed biochemistry experiments. TZ and TL generated the isogenic CRISPR-CAS9 mutant iPSC lines. TL and SG performed iPSC culture, macrophage differentiation, phagocytosis and efferocytosis assays, flow cytometry, microscopy, qPCR and ELISA experiments, and analyzed mice, with help from HY. SG, MT, MH, and TL performed and analyzed frustrated phagocytosis assays. SG, RB, and OE performed RNA-seq experiments and primary and differential analysis of the RNA-seq data. CN and SN provided essential reagents and protocols for efferocytosis assays. AV and NDS supervised RNA-sequencing. All authors contributed to the manuscript.

## Competing interest

Authors declare no competing interests.

## FIGURE SUPPLEMENTS

**Figure 1-figure supplement 1.**
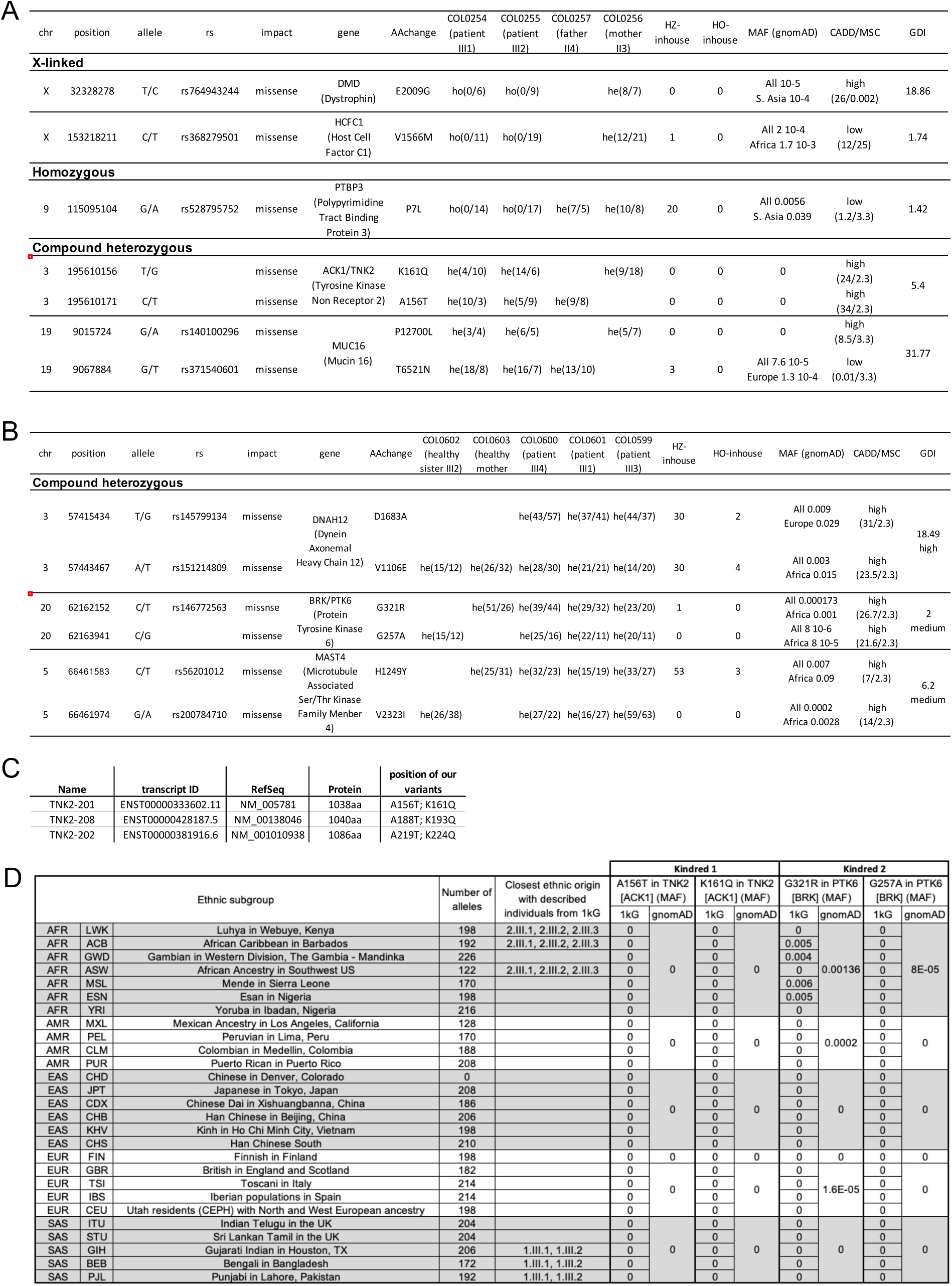
Candidate genes identified by WES analysis in family 1 and family 2. **(A) Candidate genes identified in family 1**. For a complete penetrance, 3 models of inheritance were applied: homozygous, X-linked, and compound heterozygous. Non-synonymous coding mutation with MAF< 0.01 were selected and reported for each model of inheritance. The red box shows the ACK1 mutants identified. Total MAF and maximal MAF are reported. Amino acid positions are based on transcript ENST00000333602.11. **(B) Candidate genes identified in family 2**. For a complete penetrance, 2 models of inheritance were applied: homozygous and compound heterozygous. Non-synonymous coding mutation with MAF<0.01 were selected and reported for each model of inheritance. The red box shows the PTK6 mutants identified. Total MAF and maximal MAF are reported. Amino acid positions are based on transcript ENST00000542869.2. **(C) ACK1 transcripts** with NM number reported in Ensembl. ACK1 mutant positions in the respective transcripts are reported. **(D) ACK1 and BRK mutant alleles frequency across different ethnic subgroups.** Analysis of 27 GWAS studies of SLE (https://www.gwascentral.org/) found no common variants in the close vicinity of the ACK1 (TNK2) or BRK (PTK6) genes with a p-value lower than 5.10-8, the threshold of significance for GWAS.

**Figure 1-figure supplement 2.**
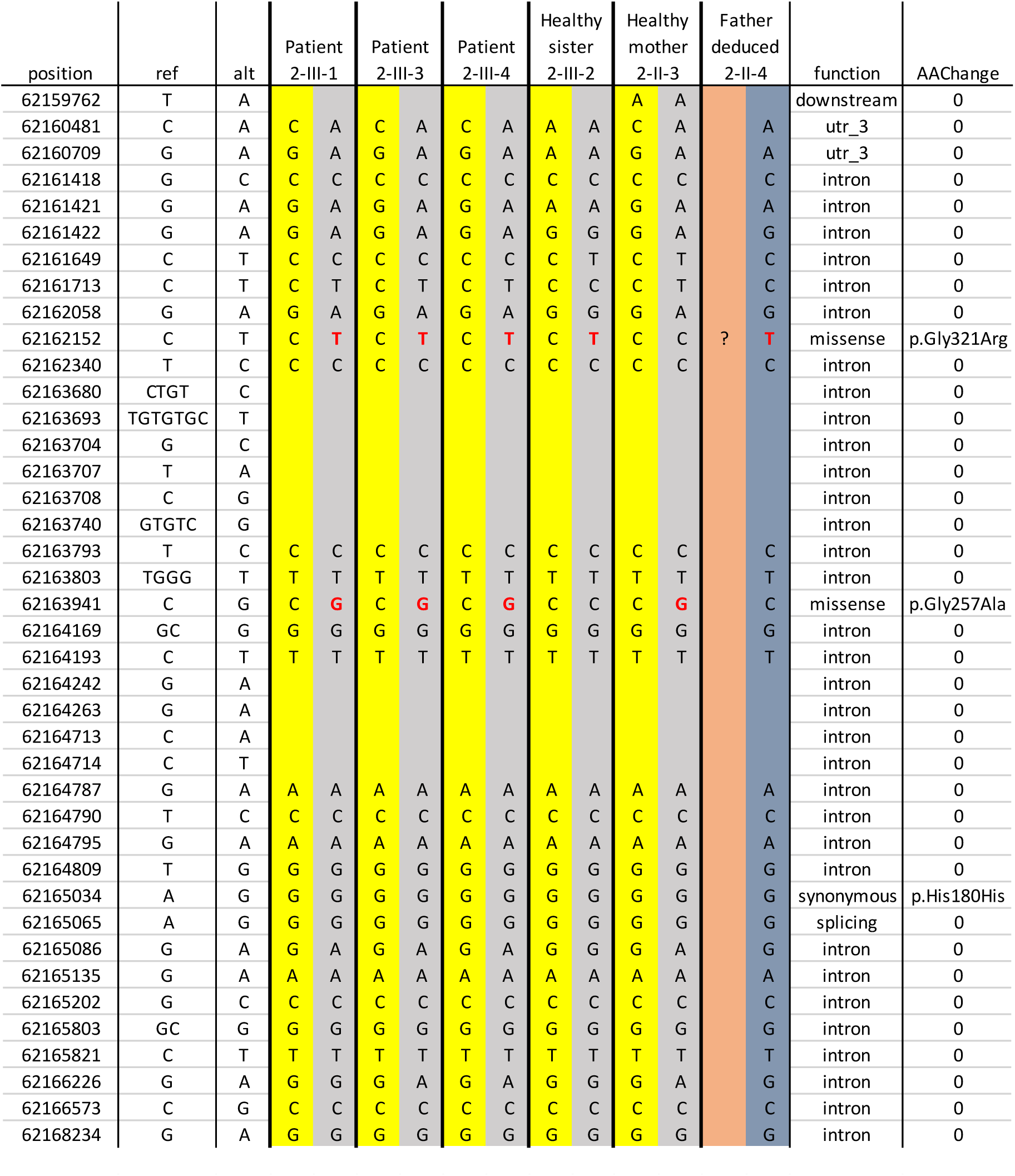
Haplotype member’s family 2. SNPs close to the G321R mutation are reported for the 3 patients, the healthy sister, the healthy mother and deduced for the healthy father.

**Figure 1-figure supplement 3.**
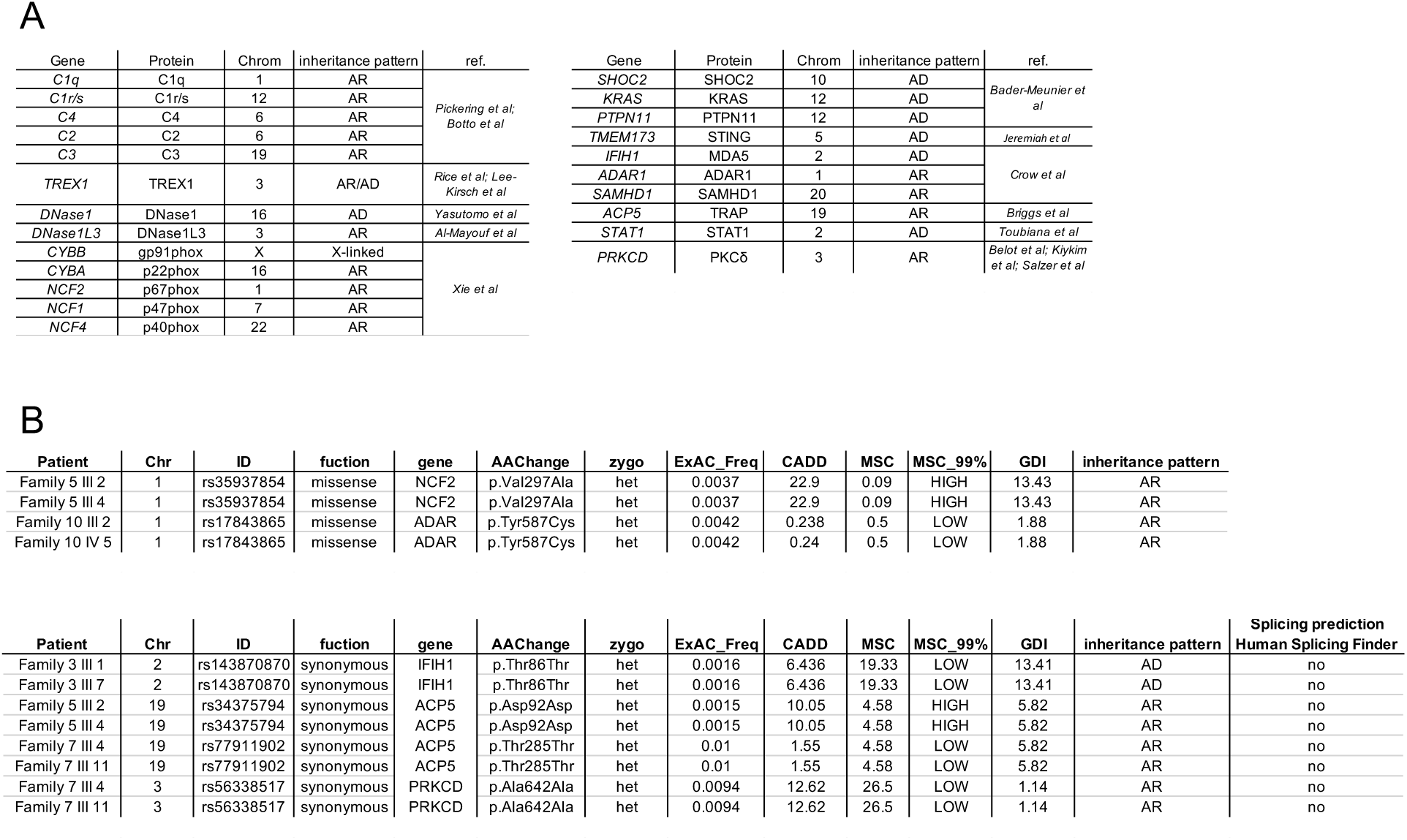
SLE-causing genes. **(A)** List of genes already described as disease-causing gene in Lupus. **(B)** Synonymous and non-synonymous mutations with a MAF<0.01 in the coding region of known SLE-causing genes identified in the 10 kindreds. Only mutations segregating with disease in the kindreds were selected.

**Figure 2-figure supplement 1.**
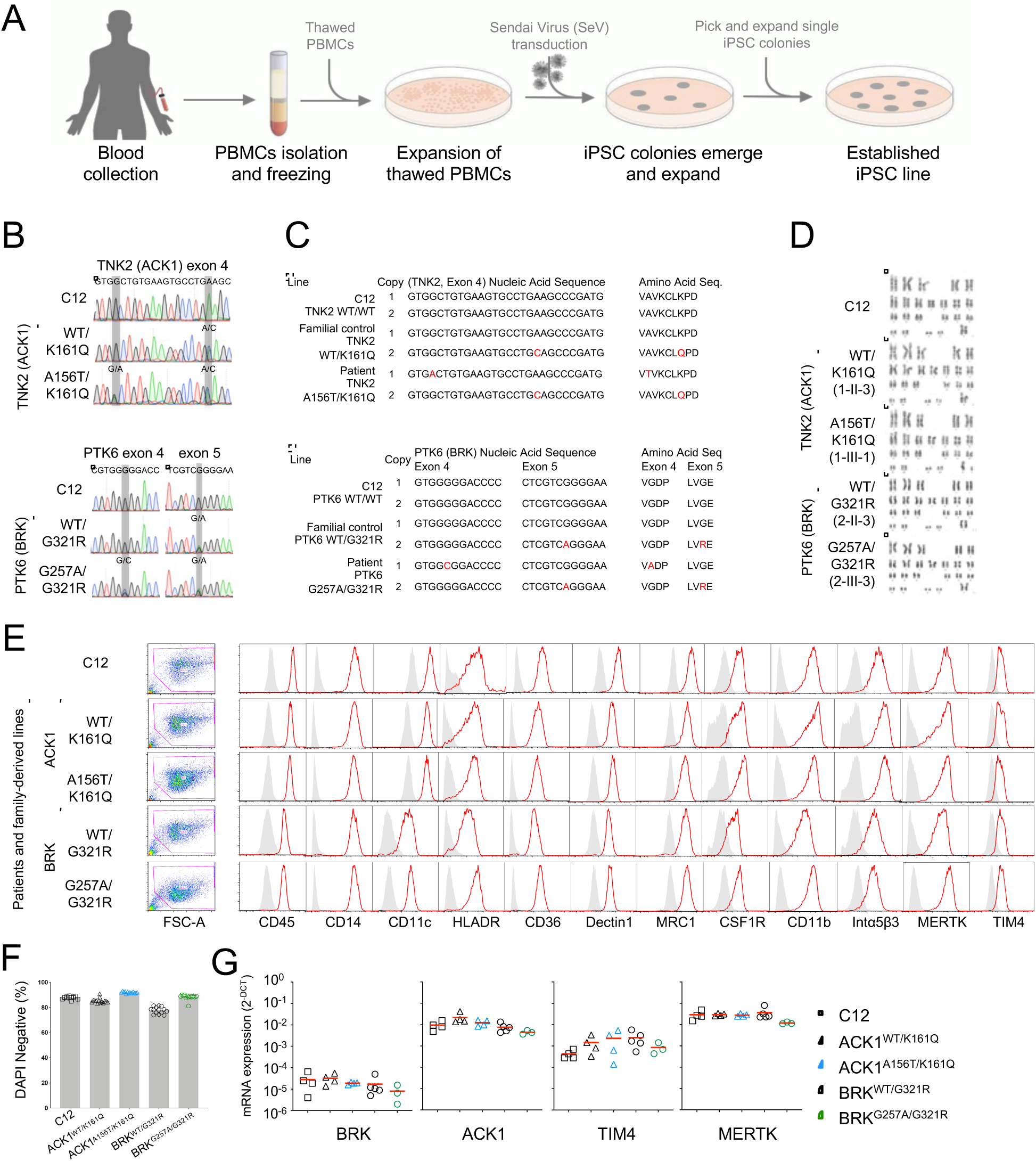
Generation and characterization of control and patient derived iPSCs and iPSC-macrophages. **(A)** Schematic representation of the process of reprograming peripheral blood mononuclear cells (PBMCs) into induced pluripotent stem cells (iPSCs). See methods for details. **(B)** Sanger sequencing of reprogrammed iPSC lines from unrelated wild type (WT) control (C12), familial controls (1-II-3 ACK1^WT/K161Q^ and 2-II-3 BRK^WT/G321R^), and SLE patients (1-III-1 ACK1^A156T/K161Q^ and 2-III-3 BRK^G257A/G321R^). Gray bars indicate positions of nucleotide substitution. **(C)** Nucleic acid and predicted amino acid sequences for both gene copies of TNK2(AKC1) or PTK6(BRK) in iPSC lines from C12 and SLE families. Variations from WT are indicated in red. **(D)** Karyotypes of iPSC lines from C12 and SLE families. Chromosome analysis was performed on a minimum of 20 DAPI-banded metaphases. **(E) Flow cytometry** analysis of surface receptor expression on iPSC derived macrophages from C12, familial controls, and SLE patients iPSC lines. Histograms show fluorescence intensity for indicated antibodies (red) and FMO controls (grey). **(F)** Bar plots show percent viability based on DAPI staining of macrophages derived from C12, familial controls, and SLE patients iPSC lines. **(G) RT-QPCR analysis of mRNA for** BRK, ACK1, TIM4, and MERTK. mRNA expression was normalized to GAPDH (2^−ΔCt^) (see methods). n≥2, from 1-2 independent experiments. P-values were obtained using an Anova test with Tukey’s correction for multiple comparisons.

**Figure 2-figure supplement 2.**
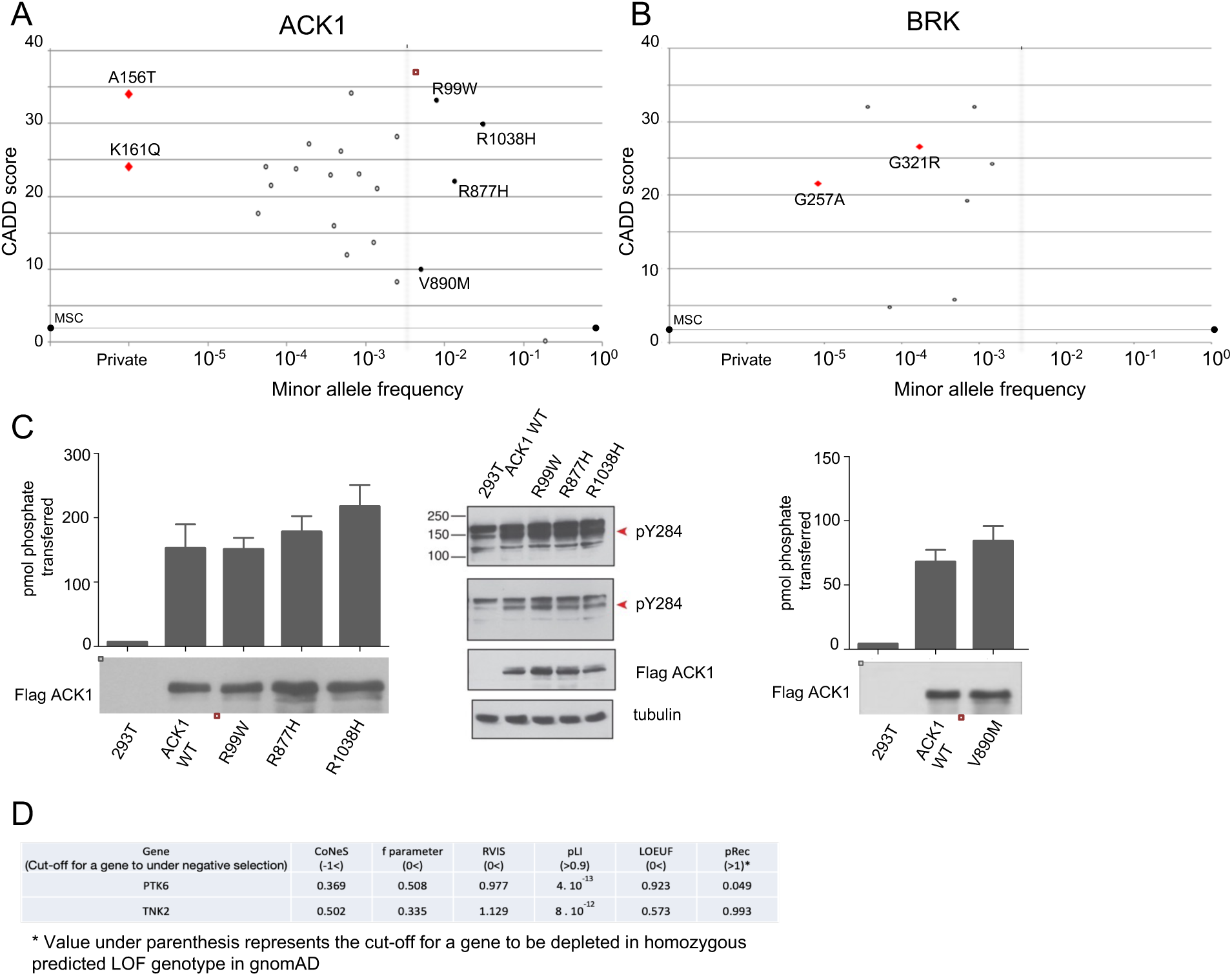
Homozygote mutant alleles reported in public database gnomAD with MAF> 0.005 don’t affect the kinase activity of ACK1. **(A, B) ACK1 and PTK6 homozygous mutants reported in gnomAD database** (A) CADD score (*y* axis) plotted against minor allele frequency (MAF, *x* axis) for the mutations found in our patients and homozygous ACK1 missense and LOF variations described in the gnomAD database. The black line corresponds to the mutation significance cutoff (MSC). ACK1^A156T^ and ACK1^K161Q^ missense variations are annotated and shown as red diamonds. Variations with MAF>0.005 (right side dotted line) and CADD/MSC high (>1, above black line) have been tested biochemically and shown as filled black circle. (B) CADD score (*y* axis) plotted against minor allele frequency (MAF, *x* axis) for the mutations found in our patients and homozygous BRK missense and LOF variations described in the gnomAD database. The black line corresponds to MSC. BRK^G257A^ and BRK^G321R^ missense mutants are annotated and shown as red diamonds. (**C**) **ACK1^R99W^, ACK1^R877H^, ACK1^R1038H^ and ACK1^V890M^ don’t impair ACK1 kinase activity.** Immunoprecipitation kinase assay (*on the le*ft). ACK1 was immunoprecipitated from HEK293T cells expressing Flag-tagged ACK1 (WT and mutants) with anti-Flag Ab. The immunoprecipitated proteins were used in duplicated in vitro for kinase reactions with WASP synthetic peptide. Western blot analysis (*on the right*). Lysates from 293T cells expressing Flag-tagged WT or mutant forms (R99W, R877H and R1038H) of ACK1 were probed with anti-ACK1 Tyr(P)^284^ (PY284), anti-Flag and anti-tubulin antibodies (Ab). **(D)** Table indicates f parameter value for negative selection in the human genome from SnIPRE ^1^, lofTool ^2^, evoTol ^3^, and CoNeS ^7^ metrics, as well as intraspecies metrics from RVIS ^4^, LOEUF ^5^, and pLI / pRec ^6^.

**Figure 3-figure supplement 1.**
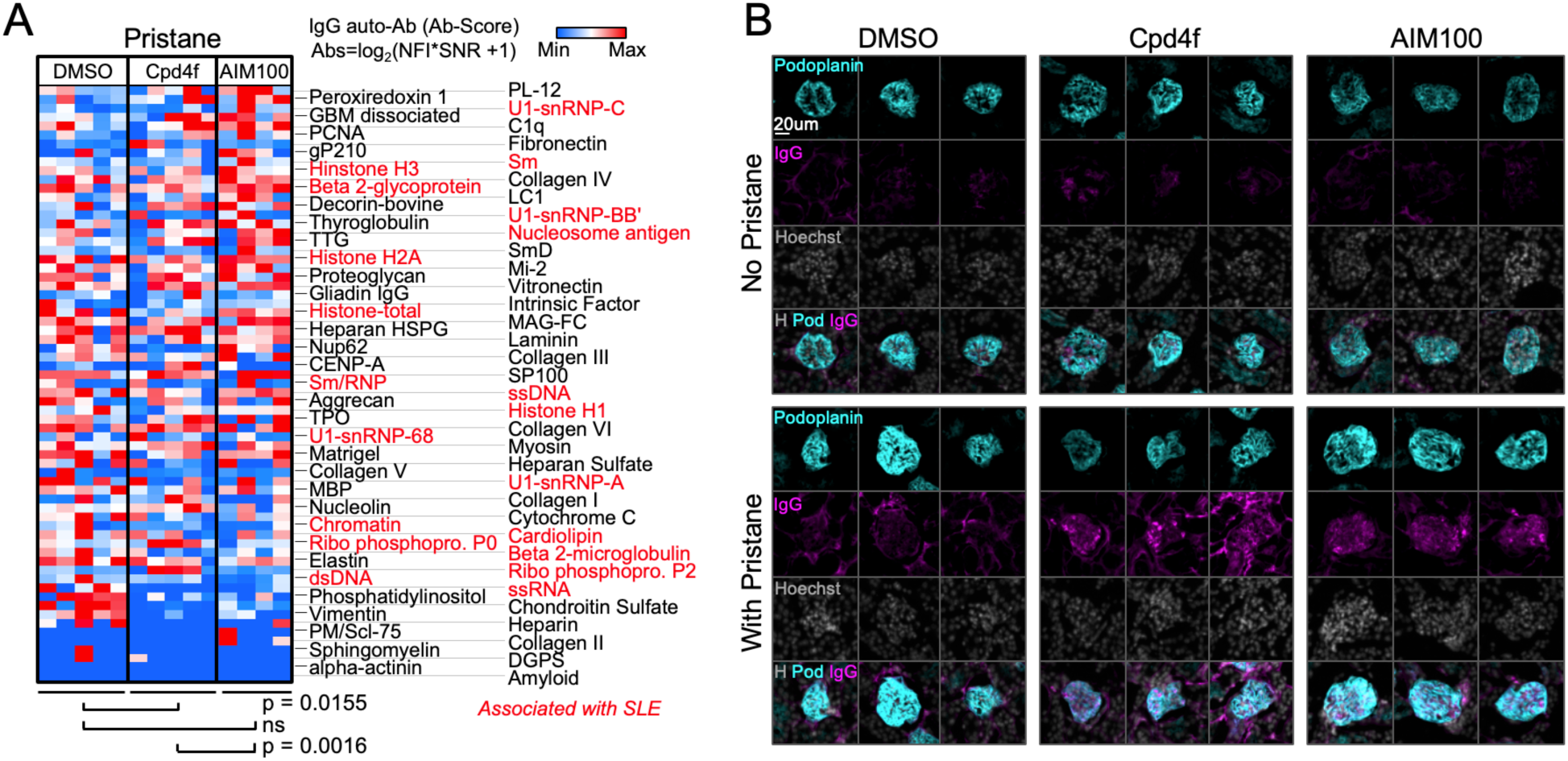
Extended analysis of ACK1 and BRK inhibitor treated mice. **(A) Heatmaps showing a comparison of serum autoantibodies detected in 4 month-old BALB/cByJ female mice.** Mice received an intraperitoneal pristane injection and a weekly intraperitoneal injection of DMSO (vehicle, 20ul/mice), AIM100 (25mg/kg in 20ul), or Cpd4f (20mg/kg in 20ul/) since the age of five weeks. Plotted values represent Ab Scores (Log_2_ [antigen net fluorescence intensity (NFI) x signal to noise ratio (SNR) +1]. Heatmap columns represent serum analysis of independent mice (n=4-5 for each of the 3 conditions). Heatmap rows sorted top to bottom starting with most significantly increased Ab Score in Cpd4f and AIM100 mice in comparison to DMSO treated mice. P-values were calculated using a Wilcoxon matched-pairs signed rank tests. **(B) Additional immunofluorescence images for mouse IgG on kidney sections.** Representative images of glomeruli displaying the average observed signal on kidney sections from 4 month-old BALB/cByJ female mice which were left untreated or received a single intraperitoneal injection of pristane, and received a weekly intraperitoneal injection of DMSO (vehicle, 20ul/mice), AIM100 (25mg/kg in 20ul), or Cpd4f (20mg/kg in 20ul/) since the age of five weeks. Kidney sections were stained with Hoechst 33342, anti-mouse IgG, and anti-mouse Podoplanin antibody. Representative images from 250 glomeruli analyzed per kidney section/mouse (>95% of all glomeruli in an entire longitudinal kidney section) where n=4 to 5 mice per condition.

**Figure 4-figure supplement 1.**
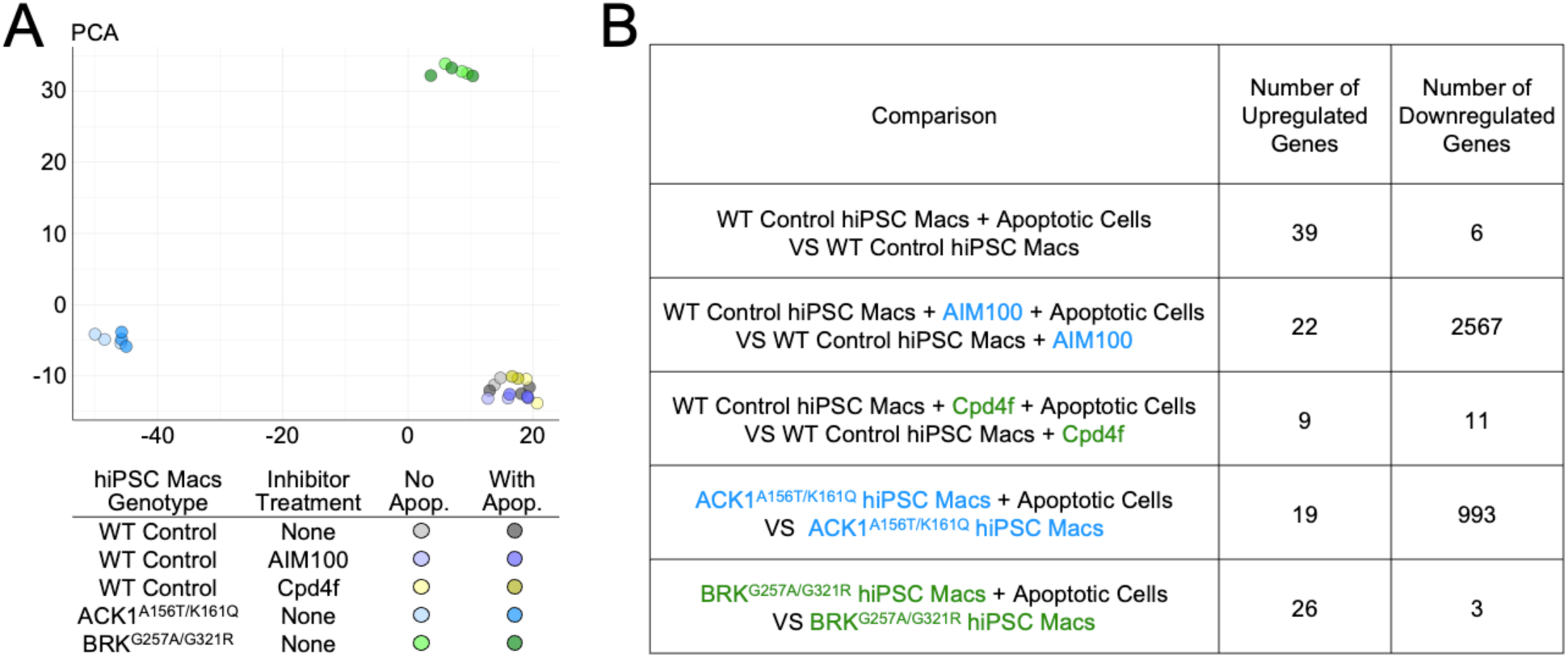
Principal component analysis (PCA) and differentially expressed genes in RNA sequencing datasets. **(A)** PCA on all samples included in the RNA sequencing dataset (treated as in figure 4E). N=2 to 3 per condition. **(B)** Table shows the number of significantly (adjusted p-value ≤0.05) upregulated and downregulated genes for specified comparisons.

**Figure 4-figure supplement 2.**
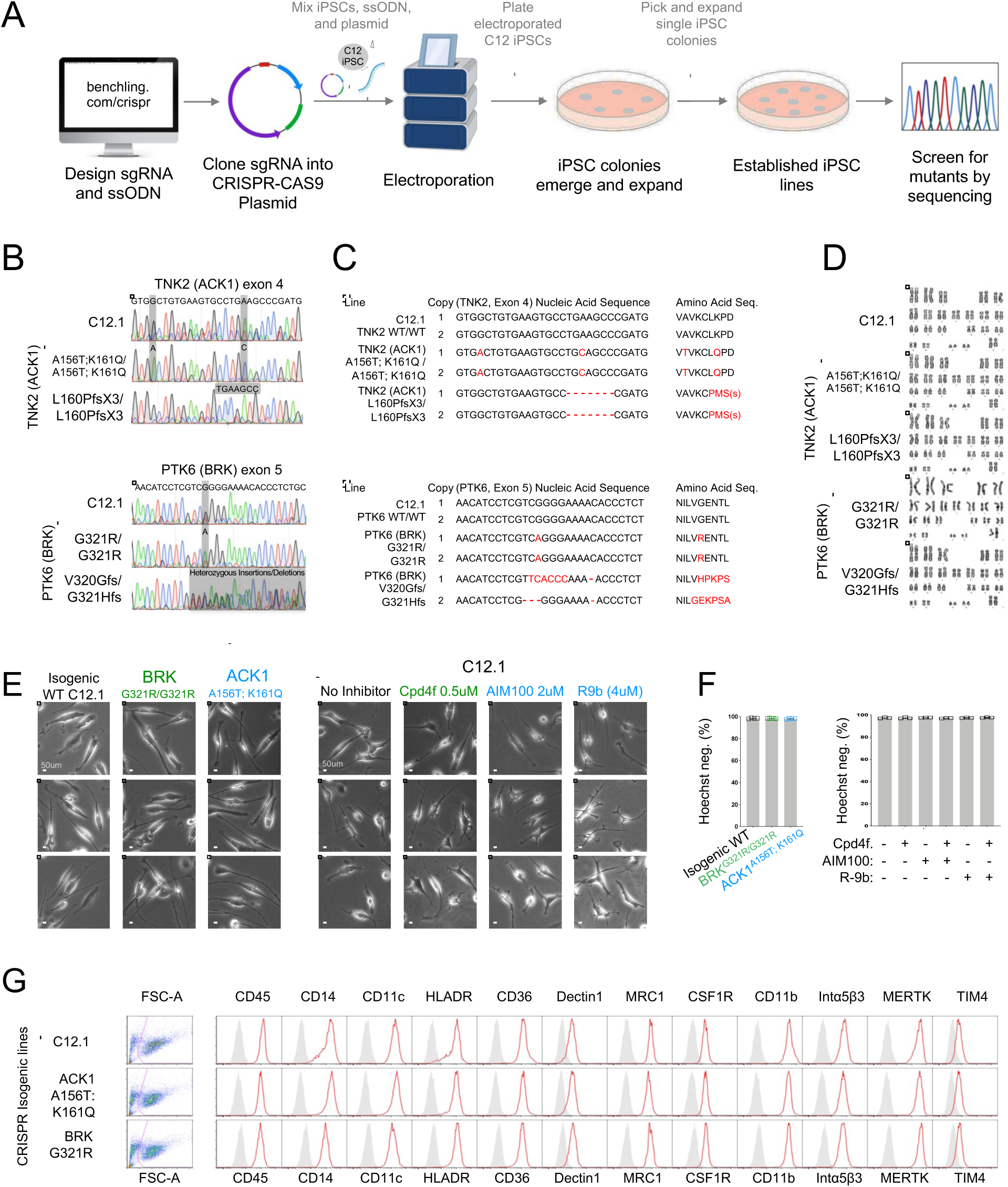
Generation and characterization of isogenic control and mutant iPSCs and iPSC-macrophages. **(A)** Schematic representation of the CRISPR-CAS9 gene editing process to derive C12 isogenic lines with TNK2 or PTK6 mutations. See methods for details. Abbreviations: single guide RNA (sgRNA); Single stranded donor oligonucleotides (ssODN). **(B)** Sanger sequencing of CRISPR-CAS9 modified isogenic iPSC lines. Gray bars indicate position of nucleotide substitutions. **(C)** Nucleic acid and predicted amino acid sequences for both gene copies of TNK2(AKC1) or PTK6(BRK) in CRISPR-CAS9 modified isogenic iPSC lines. Variations from WT are indicated in red. One dash represents one nucleotide deletion. **(D)** Karyotypes of CRISPR-CAS9 modified isogenic iPSC lines. Chromosome analysis was performed on a minimum of 20 DAPI-banded metaphases. **(E)** Representative brightfield images of Isogenic WT, BRK and ACK1 mutant macrophages (left panel) or WT macrophages treated with designated inhibitors for 2 hours (right panel). Similar morphology observed in n>5 independent experiments. **(F)** Bar plots show percent viability based on Hoechst staining of macrophages derived from isogenic iPSC lines, and percent viability of C12.1 WT macrophages after a 120 min treatment with designated inhibitors. n≥3 replicates per experimental condition. **(G) Flow cytometry** analysis of surface receptor expression on iPSC derived macrophages from isogenic WT and mutant iPSC lines. Histograms show fluorescence intensity for indicated antibodies (red) and FMO controls (grey).

**Figure 5-figure supplement 1.**
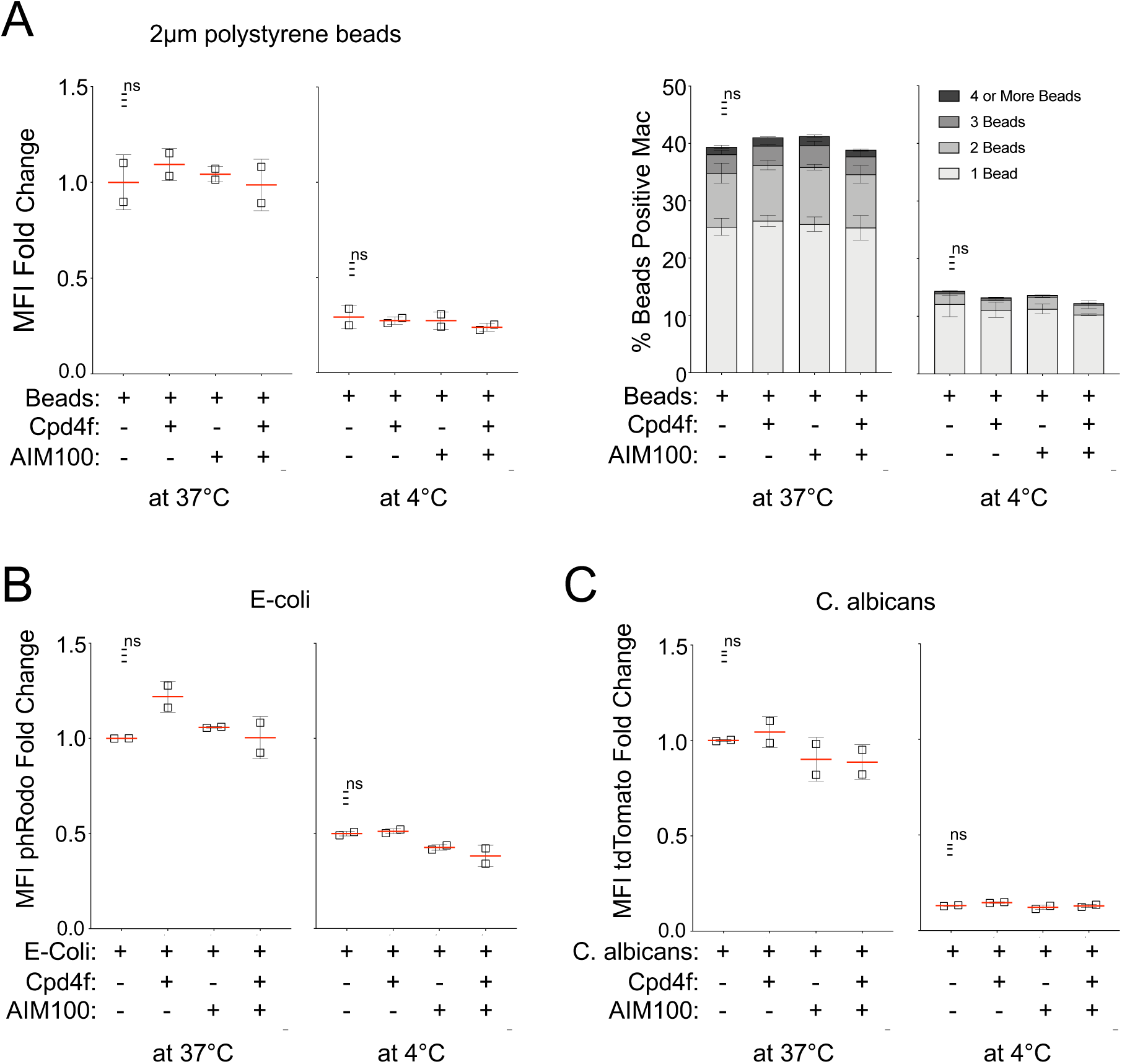
Engulfment of beads, E. coli, and C. albicans. **(A) Phagocytosis of 2µm beads.** iPSC-macrophages (C12 line) pretreated with DMSO, AIM100 (2µM), Cpd4f (0.5µM), or both, were incubated for 60 min at 37C or 4C with red fluorescent 2µm beads. Quantification of uptake is represented as fold change in mean fluorescence intensity (MFI) (670/30 nm) between DMSO treated and inhibitor treated macrophages (left panels). Quantification of uptake as percent red fluorescent (bead) positive macrophages (right panels). n=2 per experimental condition. **(B-C) Phagocytosis of E. coli and C. albicans.** Phagocytosis quantification of pHrodo labeled E. coli (B) or tdTomato fluorescent C. albicans (C) by iPSC-macrophages (C12 line) pretreated with DMSO, AIM100 (2 µM), Cpd4f (0.5 µM), or both, after a 60 min incubation at 37°C or 4°C. MFI fold change determined by dividing total MFI (585/15 nm) of individual samples by the average MFI of DMSO treated macrophages. n=2 per experimental condition. P-values in all plots were calculated using an *Anova* test (Tukey’s multiple comparisons test).

## Notes

### Competing Interest Statement

The authors have declared no competing interest.

### Author Declarations

The study was approved by the Institutional Review Board of St Thomas’ Hospital; Guy's hospital; the King's College London University and the Memorial Sloan Kettering Cancer Center. All subject samples were obtained after written informed consent from patients and their families according to the Helsinki convention (Ethics approval: 11/LO/1433). Ten multiplex families with lupus have been enrolled from Guy’s and St Thomas’ NHS Foundation Trust and UCL Hospital in London, UK from July 2010 to January 2012, following stringent criteria: a severe phenotype (lupus nephritis for at least 1 patient in each family), and a familial disease (≥2 family members affected in first degree). A total of 24 patients and 17 healthy controls from different ethnic origins (5 African ancestry, 4 Asian ancestry and 1 European ancestry) were selected. The patients each met the Systemic Lupus International Collaborating Clinics (SLICC) classification criteria for SLE2. Lupus nephritis was confirmed by a kidney biopsy classified per the 2004 ISN/RPS (International Society of Nephrology/Renal Pathology Society) classification and verified independently by 2 renal histopathologists. One hundred Mauritian participants were enrolled under the Ethical Clearance provided by the University of Mauritius Research Ethics Committee. Written consent with due signatures was recorded from all participants prior to partaking in the study. Consent was documented on a confidential form in duplicate, with one copy given to the participants for their records. The University of Mauritius Research Ethics Committee approved, sanctioned and fully endorsed this mode of consent recording. The ethnic backgrounds of the 100 Mauritian participants consisted of 26 Creole, 16 Franco-Mauritian, 21 Indo-Mauritian, 2 Sino-Mauritian, 24 other or undisclosed Mauritians.

### Summary of Updates

Figure 2 was revised and split in revised figure 2 and revised figure 3. statistics were recalculated. Methods section was updated.

